# Untangling Empowerment and Contraceptive Use Longitudinally in Five African Settings

**DOI:** 10.1101/2024.10.30.24316433

**Authors:** Marita Zimmermann, Cara Myers, Diva Dhar, Michelle O’Brien

## Abstract

**Objective:** Empowerment impacts reproductive health behaviors profoundly. Similarly, contraceptive access and use result in broader social and economic opportunities. We aimed to quantify the directionality of this cyclical relationship.

**Study design:** This study utilizes data from the Performance Monitoring for Action (PMA) surveys in Kenya, Nigeria (Kano), Burkina Faso, Democratic Republic of Congo (DRC), and Uganda. Using annual longitudinal data (2019-2021), we employed cross-lagged panel models (CLPM) to analyze the bidirectional relationship between women’s empowerment and contraceptive use.

**Results:** There was a significant positive impact of contraceptive use on women’s empowerment. In Kenya and Kano, contraceptive use was associated with a 10.1% and 12.0% increase in paid work and 14.5% and 14.8% greater control over wages, respectively. In Kenya, women doing paid work with wage control were 5.2% more likely to use contraception the following year. In Kano, control over partner’s wages increased contraceptive use by 9.6%. Additionally, having savings, financial goals, increased financial autonomy, and knowledge of financial information were associated with higher future contraceptive use.

**Conclusions:** Contraceptive use can act as a catalyst for empowerment, while empowerment, particularly decision-making autonomy over finances, enhances contraceptive use. However, these dynamics vary across different settings, emphasizing the need for context-specific strategies.

## Background

Women’s empowerment and contraceptive use are intricately linked, each influencing the other in a cyclical relationship. Empowerment, broadly defined as the process by which women gain control over their lives and decisions, has profound implications for reproductive health behaviors. Similarly, access to and use of contraception can significantly impact women’s empowerment by enabling them to make informed choices about their reproductive health, leading to broader social and economic benefits and opportunities.

A strong associational relationship between contraceptive use and empowerment has been demonstrated around the world (1–5). A systematic review found that contraceptive use among adolescent girls and young women is associated with higher education, economic opportunities, and decision-making capabilities (6). Similarly, data from Bangladesh indicated that higher levels of sexual empowerment, such as the ability to refuse sex and negotiate condom use, are significantly associated with higher contraceptive use (7). Further evidence from southeast Asia and sub-Saharan Africa underscores the multifaceted dimensions of women’s empowerment and contraceptive use (8)(9).

Although studies have consistently demonstrated a positive and significant association between contraceptive use and many aspects of empowerment, a mechanistic understanding of the causal relationship is largely missing from the current literature. Does contraceptive use empower women, or do empowered women use contraception? A few studies have been able to examine limited aspects of these causal pathways. In Malaysia, a family planning policy that combined community outreach and the distribution of contraception raised girls’ educational attainment substantially, with exposure to family planning early in life increasing the probability that women earned wages later in life (10). In Bangladesh, household delivery of family planning programming led to increased social status for women (11). In Colombia, a widespread family planning program was shown to lead to substantial socio-economic gain, despite a moderate impact on fertility outcomes (12). In one of the most studied family planning programs in Matlab, Bangladesh, results are mixed, but many indicators of the welfare of women and children have been shown to improve significantly over several decades with increased contraceptive uptake (13,14). However, results continue to be wide-ranging. In a study in Kenya, Nigeria, and Senegal, contraceptive adoption had both positive and negative effects across empowerment domains of egalitarian beliefs, reproductive control, financial autonomy, and overall autonomy, and the effects varied by setting (15). In the other causal direction, previous work has demonstrated that increased economic opportunities do lead to decreased fertility, for example in India (16) and Bangladesh (17), though contraceptive use was not directly assessed.

This multidirectional relationship is further complicated by challenges in defining the process of empowerment. We referenced the Can-Act-Resist Framework (18) (Raj, 2020) which operationalizes this process through a series of questions that identify key measures and indicators: Do you want to engage in a particular behavior? (choice and aspiration); Can you engage in the behavior? (CAN); Have you engaged in the behavior? (ACT); What happened as a consequence of this action? (consequences); and If there was backlash, did you continue to engage in the behavior? (RESIST). We consider both economic empowerment and social empowerment that may come about as a result of family planning utilization. Economic empowerment is defined as a woman’s ability to succeed and advance economically and her power to make and act on economic decisions. This includes factors such as labor force participation (employment status, hours, occupational type), education (literacy, numeracy, school enrollment, educational attainment, job training), autonomy and household decision-making power (control over household resources, labor income, time use control), and financial literacy and inclusion (savings, access to bank accounts, ability to take out loans). Social empowerment is defined as the process in which individual or social groups acquire the skills necessary for taking control of their own lives. This includes such factors as freedom of movement, voice (e.g., able to express opinions), agency (e.g., say in household decision making, how to spend free time), freedom from violence / harassment, and socioemotional wellbeing (mental health, resilience, self efficacy). Unfortunately, the existing data predominantly collects information about *economic* empowerment, thus our analysis focuses on this aspect.

The complexity of the relationships between empowerment and contraceptive use makes the causality difficult to disentangle. Likely, both directional mechanisms are occurring at different times in women’s lives, settings, and experiences, and an understanding of the dynamics of these relationships is critical for family planning and empowerment programs. One step towards building this understanding is the use of longitudinal data, which allows us to show which of these factors is changing first and impacting the others. This paper aims to utilize such data to explore the causal relationship between women’s empowerment and contraceptive use. By examining the temporal sequence and directionality of these variables, we seek to uncover when empowerment leads to increased contraceptive use, when contraceptive use serves as a catalyst for women’s empowerment, or both. Understanding this relationship is crucial for policymakers and program designers aiming to improve reproductive health outcomes and advance gender equality.

## Methods

### Data Source

For this analysis, we used data from the Performance Monitoring for Action (PMA) surveys in Kenya, Nigeria (Kano), Burkina Faso, Democratic Republic of Congo (DRC), and Uganda. PMA is a nationally representative survey completed annually. Survey participants are women aged 15 to 49 who are asked about their background, birth histories, family planning methods, fertility preferences, community norms, aspirations, intentions, empowerment, and more. The survey is based on a multi-stage cluster design with urban-rural county as strata. Households are randomly sampled within each representative geographical cluster. Beginning in 2019, PMA implemented a panel design embedded within the cross-sectional surveys, allowing for longitudinal analysis. We used three years of longitudinal data from 2019, 2020, and 2021. Selected settings had three waves of data available at the time of analysis.

To measure contraceptive use, we chose to look at current use of any method. Methods included implants, injectables, pill, IUD, condoms, sterilization, withdrawal, and other modern or traditional methods. We were also interested in separating the intent to use contraception from the ability to obtain it. To this end, we performed a secondary analysis using intent to use contraception in the next 12 months as the metric. In the PMA data, this question was only asked of women not currently using a method, therefore the secondary analysis is limited to this population.

To measure empowerment, we included several metrics (Table 1). These included survey questions measuring components of work, decision making autonomy, and financial autonomy. For the decision-making autonomy questions with answer choices of “you,” “your husband/partner,” “you and your husband/partner jointly,” or “someone else,” responses of “you” or “you and your husband/partner jointly” were coded as 1, while responses of “your husband/partner” and “someone else” were coded as 0. We also included two aggregate scale metrics, for household decision making and financial autonomy, based on a confirmatory factor analysis on the PMA data (19). While we recognize that empowerment encompasses more than just the factors listed above, we are restricted by the indicators available in the PMA dataset.

**Table 1.**
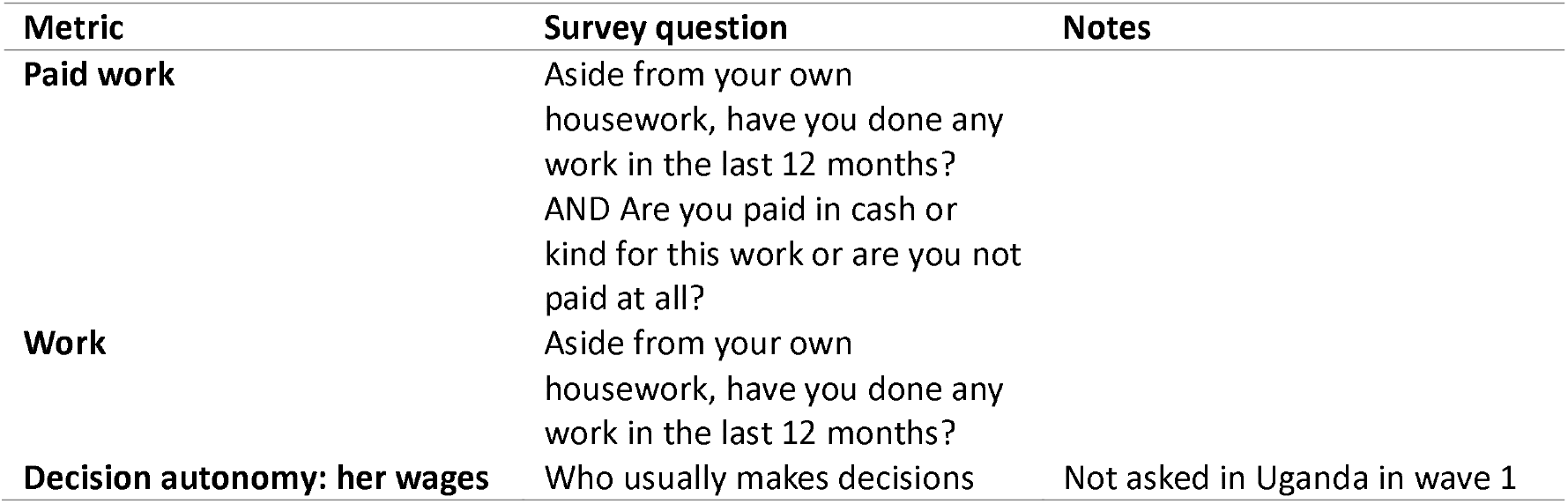
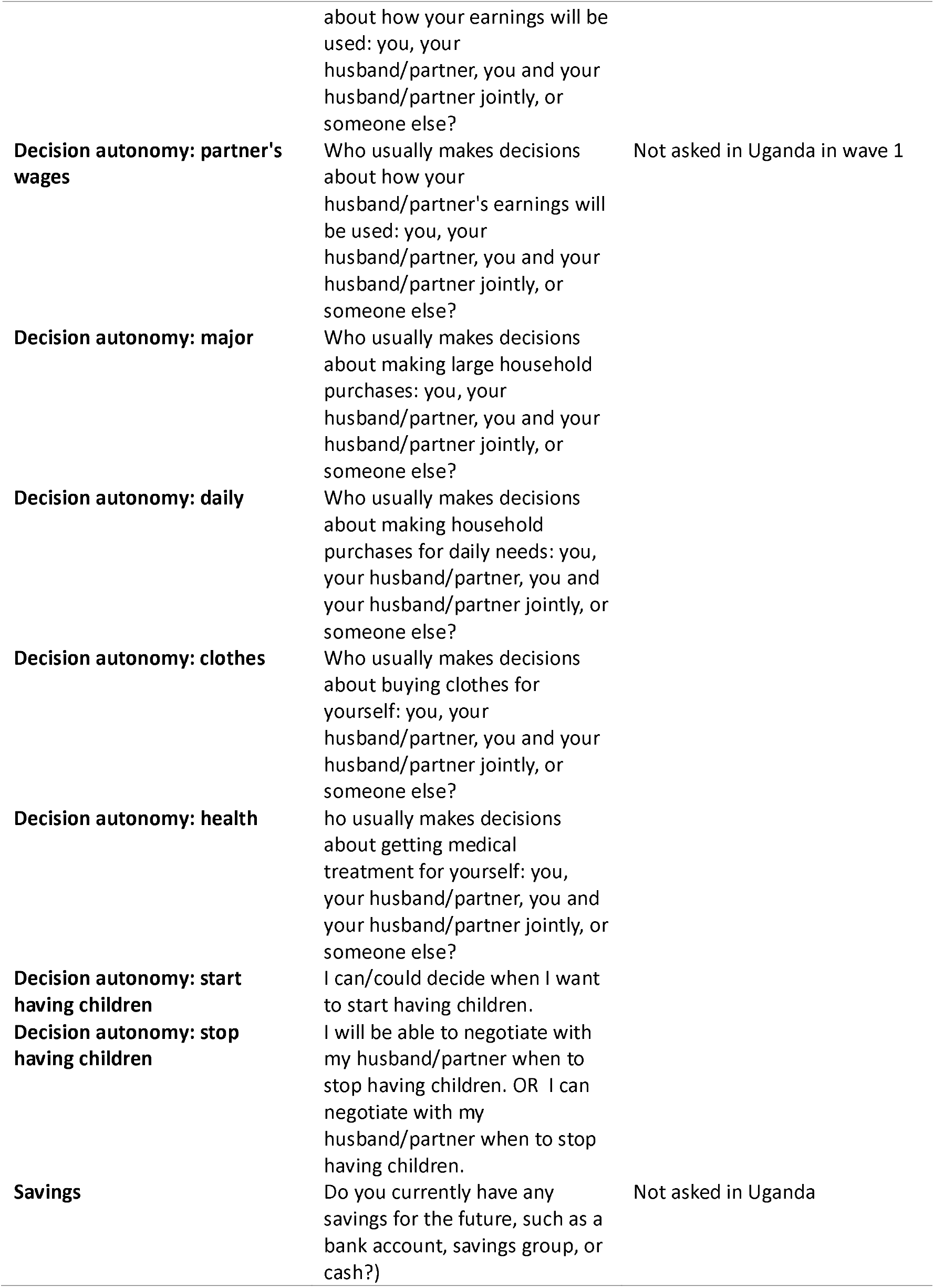
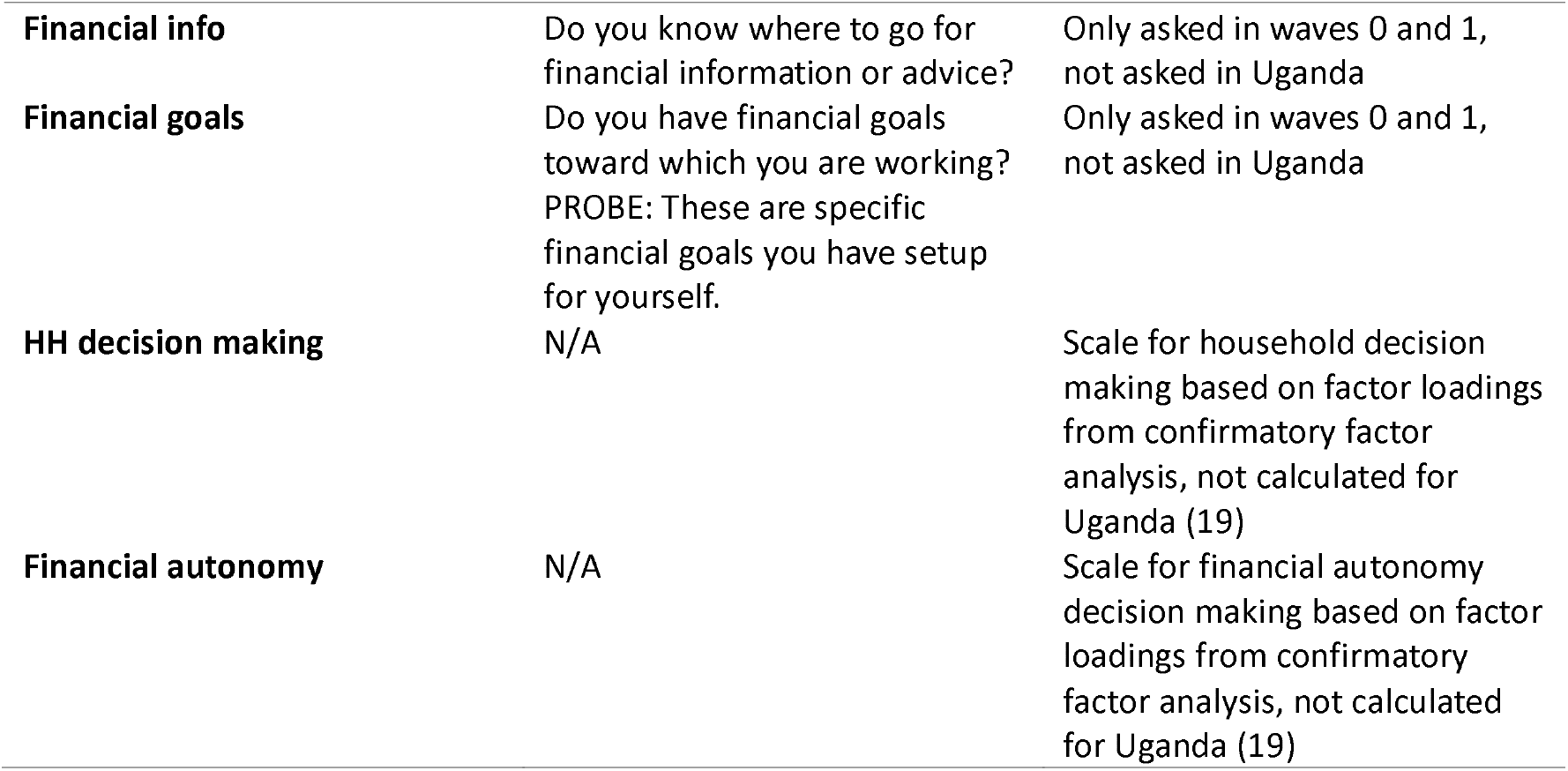
Empowerment metrics included in analysis. Source PMA data.

### Cross lagged panel models

For this analysis, we used cross-lagged panel models (CLPM). A CLPM is a type of structural equation modeling designed to measure time-lagged associations between two longitudinally assessed variables. The structure of the CLPM is shown in Figure 2. A CLPM can measure cross-lagged effects (λ), the impact of one variable at a previous timepoint on current value of another. This is the key outcome for this analysis. Additionally, it can measure stability coefficients (β), the impact of previous time on current time and time-specific correlations, i.e. the correlation between variables of interest at each time point. These are not key outcomes for our analysis but are important to control for. We also adjusted for confounders at baseline (θ), including age, marital status, highest level of education completed, number of live births, and urban/rural (expect in DRC, where the sample was only in an urban area). The primary model used was implemented as a system of equations described below, and fit as a system of linear probability models.

**Figure 1.**
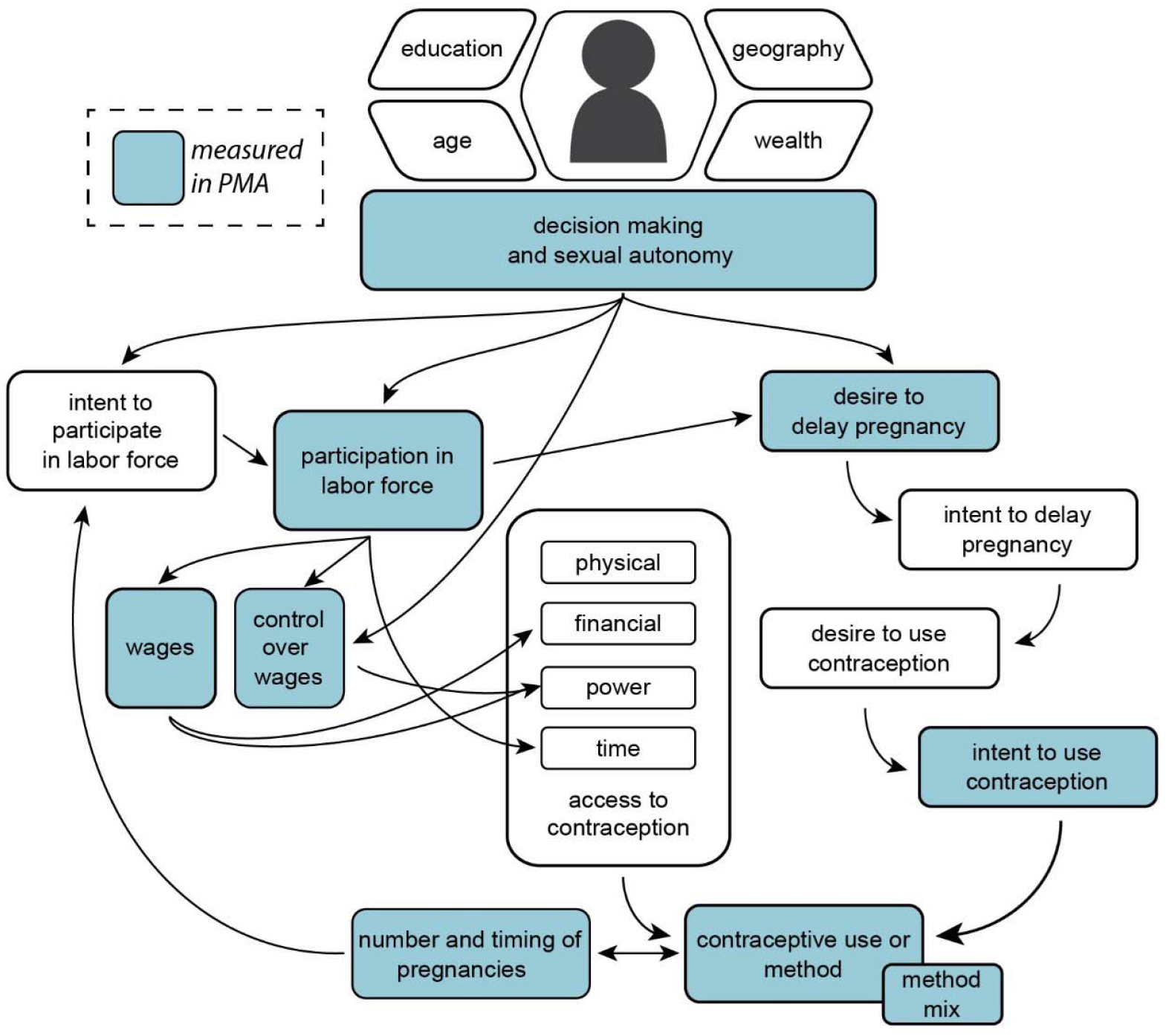
Causal framework. Several aspects of empowerment including labor force participation, control over wages, and decision making and sexual autonomy are intricately and cyclically linked to contraceptive use. The casual framework is complex, and not all aspects are currently measured in the data.

**Figure 2.**
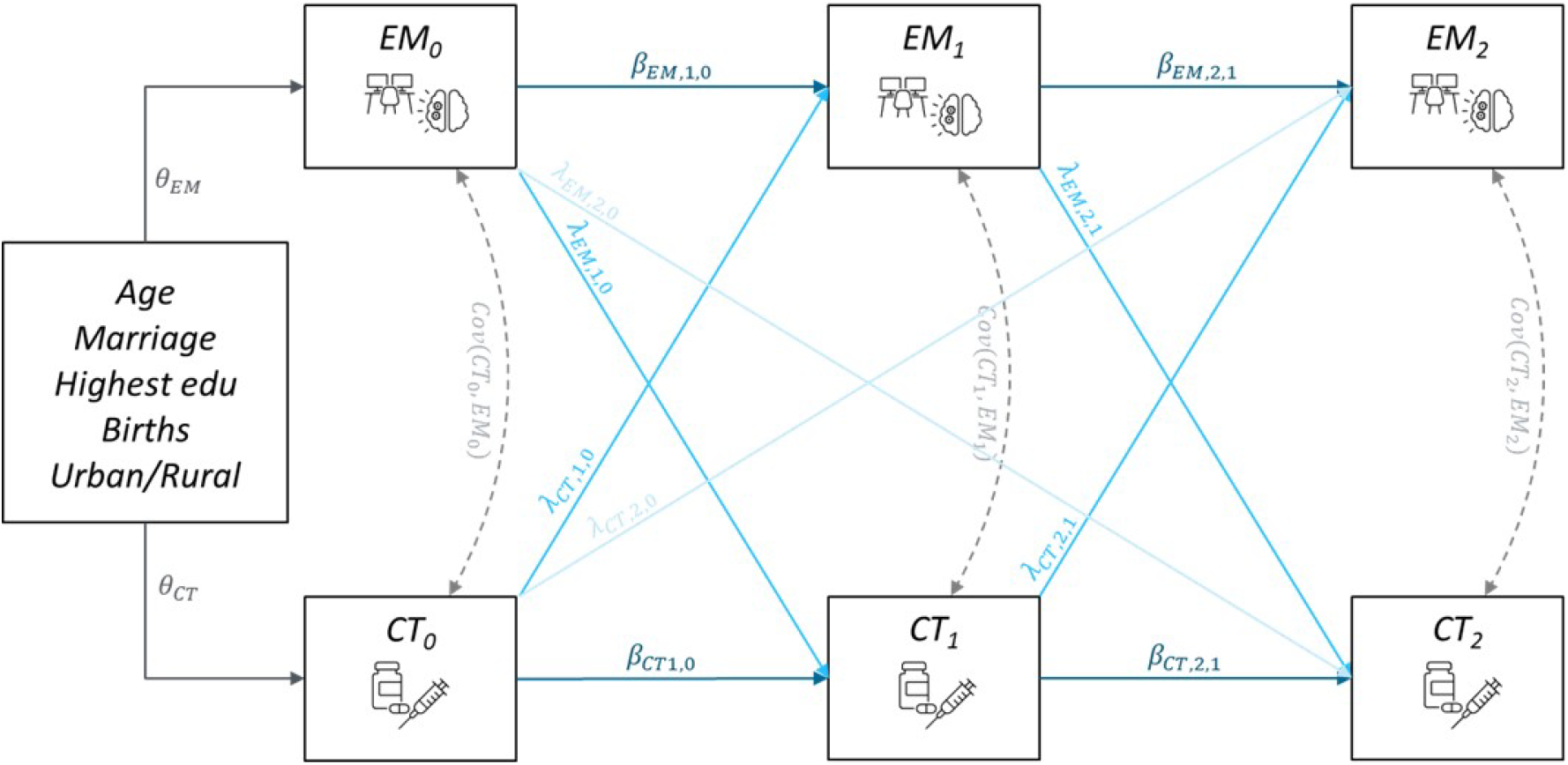
Cross lagged panel model structure, associating empowerment (EM) and contraception (CT) metrics across three time points. Blue arrows are lagged effects, dark blue arrows are stability coefficients, and dashed lines are covariances.

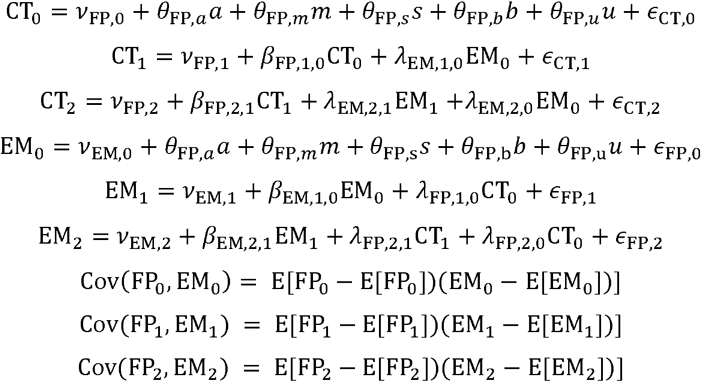

Where CT is contraceptive use, EM is each of the empowerment metrics, ν represents the intercept, β is the stability coefficients, λ is the cross-lagged effects, ⍰ is residual coefficients, θ is confounder coefficients, a is age, m is marriage, s is highest level of education, b is number of live births, and u is urban/rural.

Finally, we were interested in examining the CLPM results in more specific subpopulations. We therefore stratified all models by five separate covariates: work in the agricultural (ag) sector/non-ag sector, married/unmarried, age over/under 25, completed/did not complete primary school, and wealth quintile 1-3/4-5. These are individual stratified models, and so do not consider intersectionality and cannot be directly compared to each other.

## Results

Sample sizes ranged from 842 in Kano to 3,481 in Burkina Faso (Table 2). In Burkina Faso and Kano, over 70% of women were married, whereas less than 40% were married in DRC and Uganda. Women in Kano and Uganda had had more births. Women in Kenya had the highest contraceptive use rate (55%), and Kano had the lowest (11%), though intent to use contraception in the next 12 months was over 70% in all settings except Kano (40%). The percent of women doing paid work in the past 12 months ranged from 45% in Burkina Faso to 66% in Kano. Self-reported decision making autonomy was generally high in all settings over daily purchases and when to start and stop having children, but lower and more heterogeneous over major purchases and wages.

**Table 2.** Demographic characteristics of women in the sample by country in wave 1.

|  | Burkina | DRC | Kano | Kenya | Uganda |
| --- | --- | --- | --- | --- | --- |
| N | 3481 | 1727 | 842 | 4881 | 1506 |
| Age | 31 (0.27) | 30 (0.27) | 29 (0.28) | 31 (0.16) | 32 (0.28) |
| Married | 75% (2.7) | 32% (2.3) | 81% (2.9) | 61% (1.4) | 36% (2.5) |
| Live births | 3.6 (0.1) | 2.4 (0.12) | 4.1 (0.17) | 3 (0.053) | 4.1 (0.11) |
| School: primary+ | 20% (1.5) | 79% (2.9) | 34% (5.3) | 47% (1.3) | 32% (2.3) |
| Wealth quintile: 4+ | 38% (3.9) | 46% (4.2) | 43% (5.5) | 32% (1.4) | 38% (3.6) |
| Urban | 21% (1.8) | -- | 38% (6.7) | 25% (1.3) | 17% (2.4) |
| Intent to use contraception (12 mo) | 76% (2) | 75% (1.7) | 40% (4.6) | 82% (0.88) | 81% (1.6) |
| Currently using contraception | 35% (2) | 46% (2.8) | 11% (2.6) | 55% (0.98) | 46% (1.7) |
| Paid work | 45% (3.9) | 48% (3.8) | 66% (3.6) | 55% (1.5) | 50% (3) |
| Work | 59% (4.5) | 52% (3.8) | 66% (3.5) | 61% (1.7) | 66% (2.9) |
| Decision autonomy: major | 40% (2.9) | 64% (4) | 32% (7.1) | 74% (1.7) | 53% (2.7) |
| Decision autonomy: daily | 74% (2.7) | 76% (4.3) | 49% (9.8) | 85% (1.1) | 71% (2.6) |
| Decision autonomy: clothes | 79% (2) | 73% (4.3) | 41% (7.9) | 87% (1.1) | 74% (2.4) |
| Decision autonomy: health | 36% (2.5) | 46% (3.6) | 35% (8.2) | 78% (1.6) | 49% (2.8) |
| Decision autonomy: her wages | 35% (3.7) | 26% (2.6) | 56% (5) | 44% (1.6) | 38% (3) |
| Decision autonomy: partner's wages | 23% (2.4) | 61% (3.4) | 16% (4.4) | 55% (2) | 36% (2) |
| Decision autonomy: start having children | 67% (3.5) | 66% (3.2) | 53% (9.1) | 90% (1.2) | 80% (2) |
| Decision autonomy: stop having children | 79% (2.8) | 70% (3.7) | 53% (5.3) | 89% (1) | 83% (1.6) |
| Savings | 15% (2.1) | 18% (2.6) | 39% (6.9) | 44% (1.7) | -- |
| Financial info | 21% (2.3) | 17% (2.7) | 65% (8.8) | 50% (2) | -- |
| Financial goals | 74% (3.6) | 56% (4.8) | 88% (4.1) | 77% (1.7) | -- |

Full results of the CLPM coefficients are shown in the appendix table. In most settings, women who were economically empowered tended to be more likely to use contraception the following year, though which types of economic empowerment were relevant varied by setting. In Kano, Burkina Faso, and Kenya, women who had decision making autonomy over when to stop having children at baseline had a 7.0%, 7.5%, and 9.0% increased probability of using contraception the following year, respectively (Figure 3). In Kenya, we found that women doing paid work at baseline had a 4.4% increased probability of contraceptive use following a year, and a 5.2% increase when she also had decision making autonomy over those wages. In Kano, decision making autonomy over her partner’s wages at baseline led to a 9.6% increased probability of contraceptive use the following year. In both Kenya and Kano, having savings (5.5%, 7.4%, respectively), financial goals (9.0%, 4.6%), and an increased financial autonomy score (5.1%, 4.1%) as well as knowing where to get financial information in Kenya alone (4.7%), were also associated with a probability of using contraception one year later.

**Figure 3.**
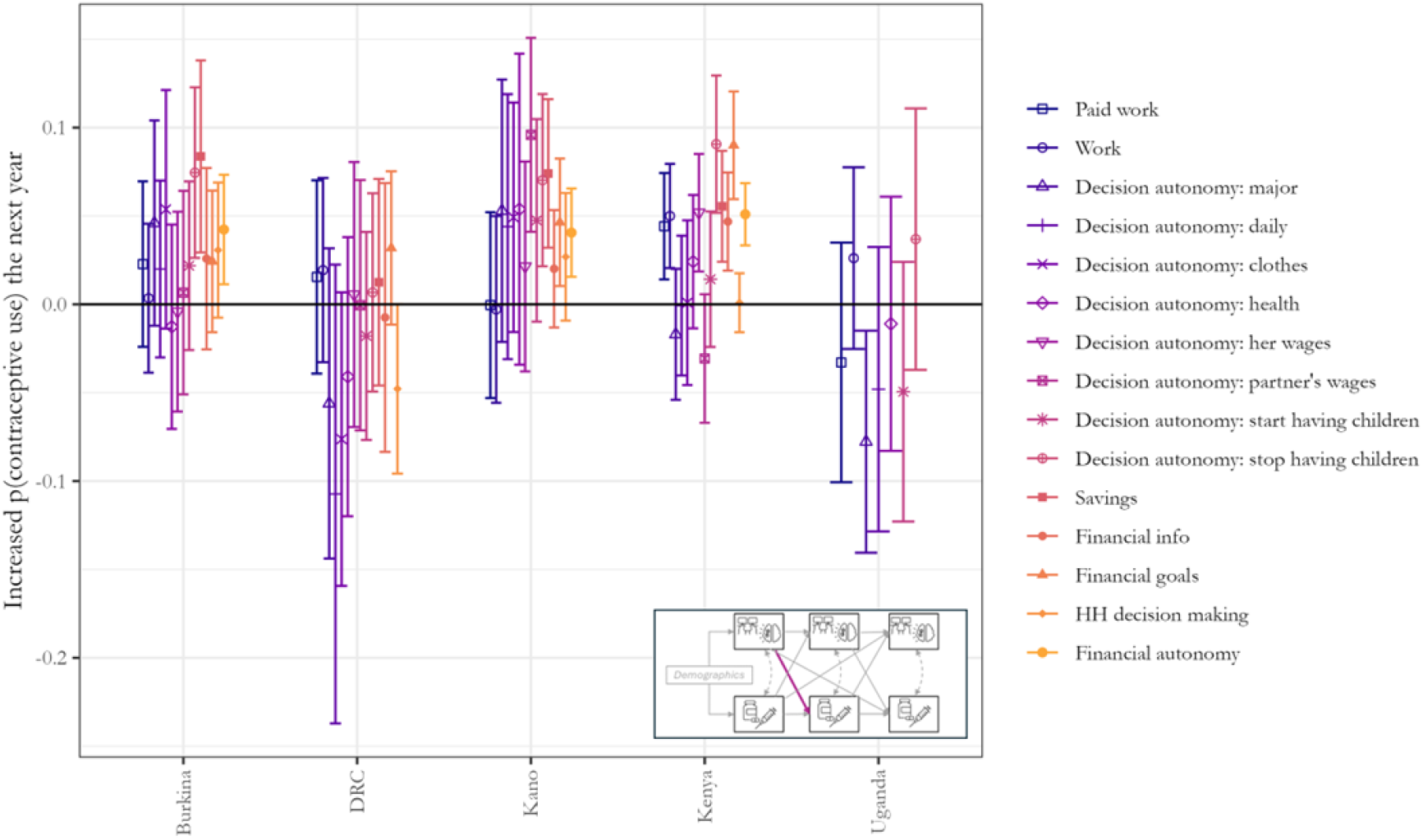
Increased probability of using contraception in year one if a woman was empowered at baseline according to one of the empowerment metrics (paid work or decision making autonomy) compared to if she was not empowered. Colors represent each empowerment metric.

In most settings, women who used contraception were more likely to be empowered the following year, though which types of empowerment they achieved varied by setting. Contraceptive use at baseline led to a significant increase in probability of having decision making autonomy over when to stop having children in year 1 in Kenya (3.7%), Uganda (4.6%), Burkina Faso (7.0%), and Kano (13.8%) (Figure 4). Contraceptive use at baseline led to an increased probability of doing paid work in Kenya (10.1%) and Kano (12.0%), as well as an even higher increase (14.5% and 14.8%, respectively), of having decision making autonomy over those wages and an increased probability of having savings (6.8%, 18.4%). In Burkina Faso, contraceptive use also led to an increased probability of having savings (6.9%) and an increase in the financial autonomy scale (9.3). In Kano, contraceptive use led to a 20% increase in decision making autonomy over major purchases. In Kenya, using contraception also led to a 5.9% increase in knowing where to get financial information, 5.9% increase in having financial goals, and a 9.3 increase in the financial autonomy scale.

**Figure 4.**
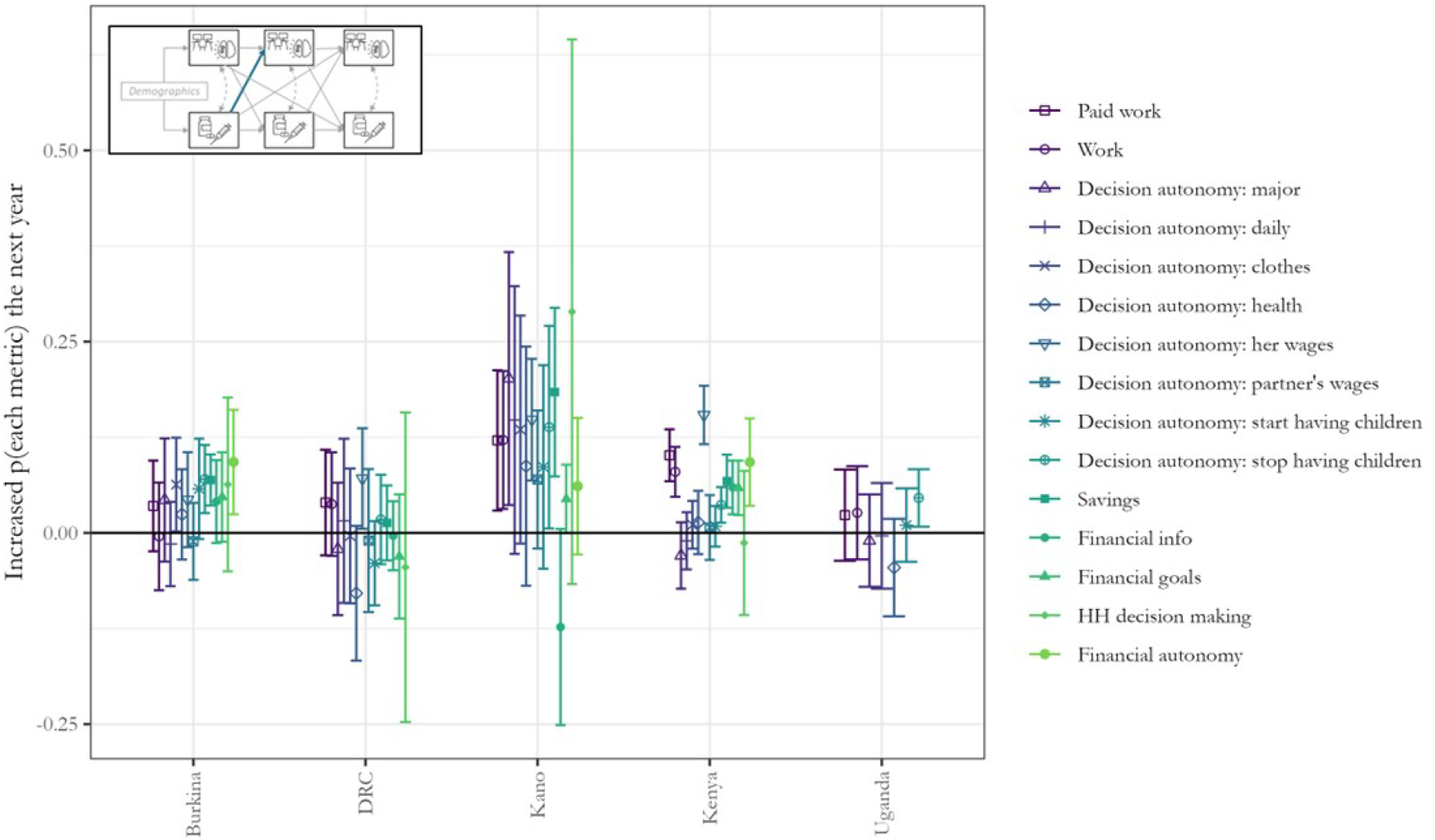
Increased probability of becoming empowered according to one of the empowerment metrics (paid work or decision making autonomy) at year one if a woman was using contraception at baseline, compared to a women not using contraception. Colors represent each empowerment metric.

Results for year 1 to 2 were similar to years 0 to 1 (appendix table). For year 0 to 2, we found few significant results. The model could not converge for financial autonomy in DRC, likely due to homogeneity within the sample population.

We found that most measures of empowerment did not increase the probability of having intent to use contraception the following year (Figure 5). In DRC and Kano, having decision making autonomy over when to stop having children was associated with a 7.2% and 17.5% increase in having intent to use contraception the following year, whereas in Burkina Faso decision autonomy over when to start having children had a higher association (6.7%). Conversely, in DRC, having control over her own wages led to a 15.6% decreased probability of having intent to use the following year, as did all of the economic empowerment related factors in Kenya, ranging from 20.4% decrease for women who have savings to a 2.8% decrease for knowing where to get financial information. In the other direction, we found that having intent to use contraception had limited associations with increased empowerment the following year. Having an intent to use contraception in the next 12 months led to a decreased probability or working in Kenya (12.3%), Kano (7.1%), and DRC (11.7%). Intent also led to a 9.4% decreased probability of having decision autonomy over her wages and 7.7% decreased probability of having savings in Kenya.

**Figure 5.**
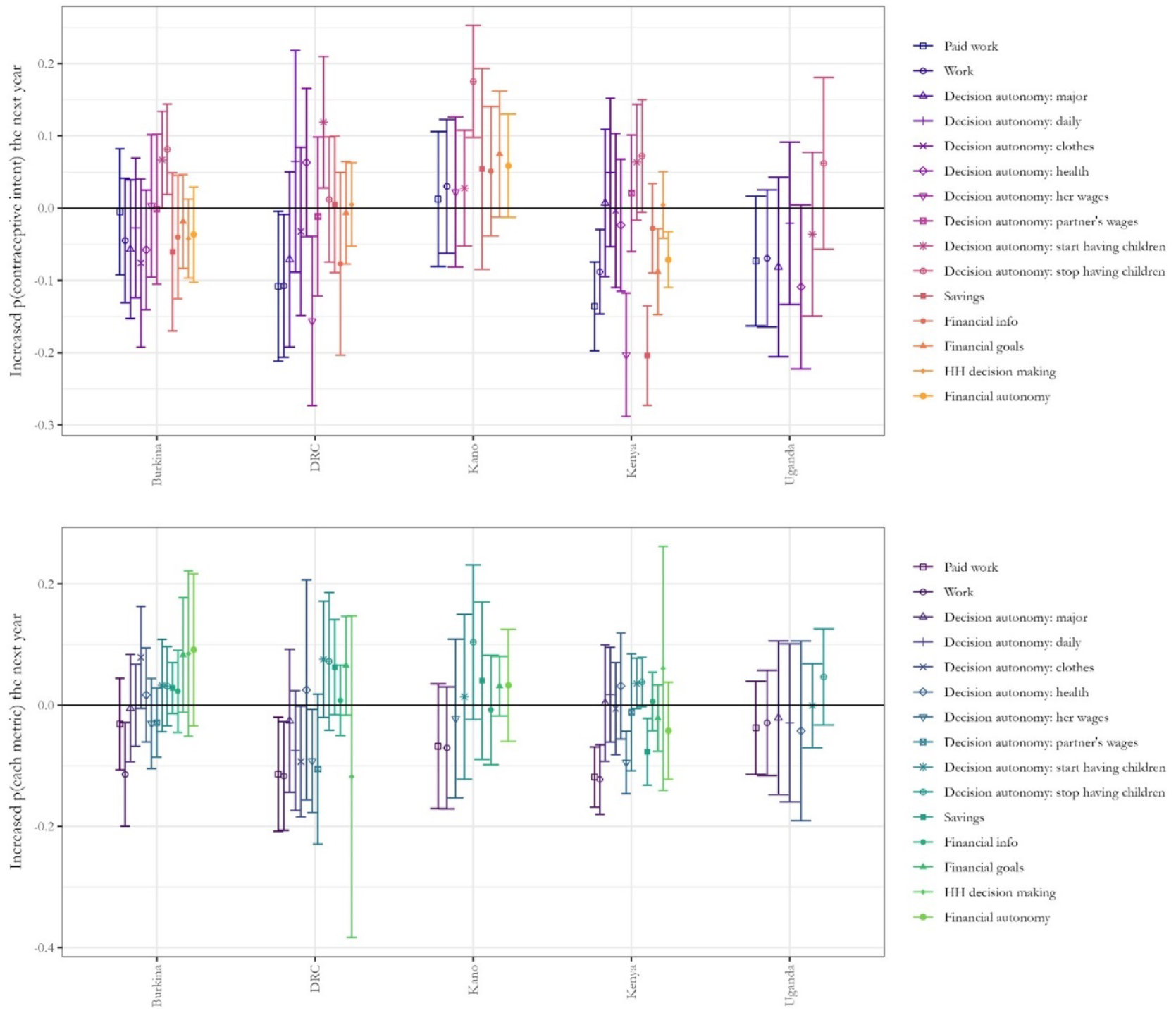
Model results using intent to use contraception among women not currently using as the contraceptive metric. Top panel shows the increased probability of having intent to use contraception in year one if a woman was empowered at baseline according to one of the empowerment metrics (paid work or decision making autonomy) compared to if she was not empowered. Bottom panel shows increased probability of becoming empowered according to one of the empowerment metrics (paid work or decision making autonomy) at year one if a woman had intent to use contraception at baseline, compared to a women who did not have intent. Colors represent each empowerment metric.

The relationship between contraceptive use and empowerment changes in subgroups by age, sector of work (agricultural or other), marriage status, education, and wealth. When we stratified the models by each of these factors, we found quite different results in each of these subpopulations. For example, in Kenya, contraceptive use led to an increase in doing paid work the following year for women under age 25 and married women, but not for those over age 25 or unmarried (Figure 6). In the reverse direction, women who were doing paid work who did not work in agriculture, were under age 25, unmarried, had higher education, or higher wealth levels, were more likely to use contraception the following year, whereas women who worked in agriculture, were under age 25, married, had lower education or lower wealth, were not more likely to start using contraception. Alternatively, in Burkina Faso, only women with higher education were more likely to do paid work the following year, and only women doing paid work with higher education were more likely to use contraception the following year. Which subpopulations showed significant effects varied for each metric (Appendix figure).

**Figure 6.**
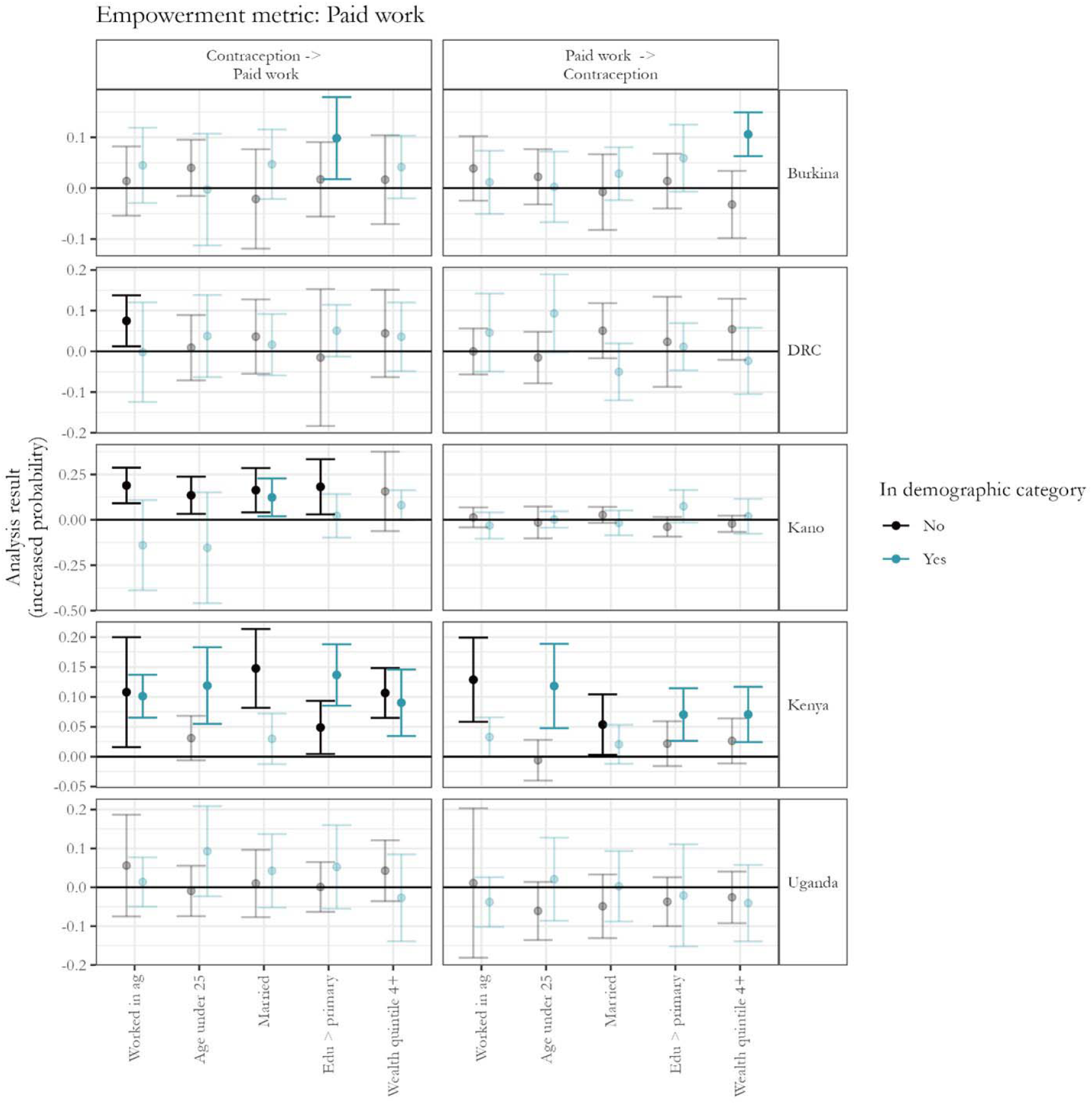
Results of the stratified CLPMs for paid work (other metrics in appndix). The points represent the increased probability of having an empowerment metric the year following using contracetion compared to not using contracption (left) and the increased probability of contraceptive use the year following having and empowerment metric compared to not having that metric (right). Each empowerment metric is a horozontal panel. Models are stratified by the five factors on the x-axis, with the blue point representing results for whome who do have that characteristic, and the black point representing women who do not have it. Statistically significant results are in brighter colors, and insignificant results are faded.

## Discussion

The findings from this study underscore the intricate and bidirectional relationship between women’s empowerment and contraceptive use. Utilizing longitudinal data from the Performance Monitoring for Action (PMA) surveys across five diverse settings—Kenya, Kano (Nigeria), Burkina Faso, DRC, and Uganda—we provide valuable insights into how these variables influence each other over time.

Our results demonstrate that contraceptive use can act as a catalyst for various aspects of women’s empowerment. For instance, contraceptive use significantly increased the likelihood of decision-making autonomy over reproductive choices in most settings. This suggests that by enabling women to control their fertility, contraceptive use may empower them to assert greater autonomy in other areas of their lives, such as more flexibility to work outside the home, pursuit of higher education, choice of relationships, leadership roles, or civic engagement. For example, in Kenya and Kano, contraceptive use was associated with increased participation in paid work and greater control over wages. This finding suggests that in some settings, contraception not only allows women to space and limit births (which reduces the physical and time constraints associated with childbearing), but also enhances their ability to engage in economic activities and to gain financial autonomy.

Conversely, the study also reveals that women’s empowerment, particularly decision-making autonomy over reproductive choices, significantly enhances the likelihood of contraceptive use. This finding aligns with previous research indicating that empowered women, who have greater control over their reproductive decisions, are more proactive in using contraception to manage their fertility and health outcomes. Furthermore, the data from Kenya highlight that economic empowerment, as indicated by engagement in paid work and autonomy over wages, also contributes to higher contraceptive use, though this finding was inconsistent across settings. This reinforces the notion that economic empowerment and reproductive autonomy are closely linked, as financial independence can provide women with the resources and confidence to make informed reproductive choices.

Interestingly, the relationship between empowerment factors and contraceptive use was quite different from the relationships with contraceptive intentions. While we see the hypothesized effect of empowerment leading to increased contraceptive use, and vice-versa, in many settings, we do not see a consistent impact on intent to use. Economic empowerment factors, such as control over wages and access to savings, were associated with a decreased future intent to use contraception in both DRC and Kenya, and having intent to use contraception led to a decreased probability of working in the future. While this finding may indicate an inverse relationship between intent to use and economic empowerment, it is likely that women who are economically empowered and intend to use contraception have more access, therefore have become users rather than continuing to express intent alone. This is an important differentiation between intent to use, which has strong societal, personal, and familiar influences, and access to contraception, which we found to be more directly impacted by economic empowerment in some settings.

The variations observed across different settings underscore the importance of context-specific strategies. For instance, the high decision-making autonomy over reproductive choices in Kano leading to increased contraceptive use contrasts with the relatively lower rates of paid work among women in the same region. This indicates that while reproductive autonomy is crucial, economic empowerment initiatives might need to be tailored differently across contexts to effectively enhance contraceptive use and overall empowerment. While there are general trends, the specific socio-cultural and economic contexts play a critical role in shaping these dynamics. This conclusion is supported by the stratified results, which vary greatly by country, subpopulation, and the combination of both. This gives us a window of understanding into how subpopulation targeting is necessary to improve the efficacy of programs and interventions.

This study contributes to the mechanistic understanding of the relationship between women’s empowerment and contraceptive use by using longitudinal data and employing cross-lagged panel models to examine bidirectional relationships. However, the study was limited by the time scale of data. The PMA data only covers three years, which is not long enough to observe meaningful change in empowerment at a societal scale. Additionally, causal interpretations based on this study should be drawn cautiously. Significant complexity exists, and many societal and individual factors, such as societal norms and fertility preferences, are changing simultaneously with the metrics studied here. Future research should continue to explore these relationships using extended longitudinal data with additional contextual factors.

Understanding the bidirectional relationship between women’s empowerment and contraceptive use is crucial for designing effective interventions. Policies and programs aimed at enhancing women’s reproductive health need to simultaneously address various empowerment dimensions, including economic and decision-making autonomy and access to safe and effective contraception. By fostering environments where women are both empowered and have access to contraception, we can achieve broader health, social, and economic benefits, ultimately advancing gender equality and improving overall well-being. In conclusion, this study highlights the cyclical nature of empowerment and contraceptive use and emphasizes the need for integrated approaches that consider the multifaceted and context-specific factors influencing these dynamics.

## Data Availability

All data produced in the present study are publicly available.

