## appendix table for "Untangling Empowerment and Contraceptive Use Longitudinally in Five African Settings"

| Country | Empowerm | Coefficient | Estimate | SE |
|---|---|---|---|---|
| Kenya | Paid work | vEM3 | 0.228 | 0.013 |
| Kenya | Paid work | βEM,2,1 | 0.53 | 0.016 |
| Kenya | Paid work | λEM,2,1 | 0.05 | 0.012 |
| Kenya | Paid work | λEM,2,1 | 0.018 | 0.017 |
| Kenya | Paid work | vEM2 | 0.251 | 0.012 |
| Kenya | Paid work | βEM,1,0 | 0.548 | 0.015 |
| Kenya | Paid work | λEM,1,0 | 0.044 | 0.015 |
| Kenya | Paid work | vCT1 | 0.162 | 0.038 |
| Kenya | Paid work | θCT,age | -0.001 | 0.001 |
| Kenya | Paid work | θCT,marria | 0.336 | 0.022 |
| Kenya | Paid work | θCT,highes | 0.038 | 0.008 |
| Kenya | Paid work | θCT,births | 0.031 | 0.006 |
| Kenya | Paid work | θCT,urban | 0.101 | 0.024 |
| Kenya | Paid work | vEM3 | 0.254 | 0.016 |
| Kenya | Paid work | λEM,2,1 | 0.139 | 0.018 |
| Kenya | Paid work | βEM,2,1 | 0.436 | 0.019 |
| Kenya | Paid work | λEM,2,1 | 0.02 | 0.019 |
| Kenya | Paid work | vEM2 | 0.342 | 0.017 |
| Kenya | Paid work | λEM,1,0 | 0.102 | 0.017 |
| Kenya | Paid work | βEM,1,0 | 0.38 | 0.019 |
| Kenya | Paid work | vEM1 | -0.085 | 0.04 |
| Kenya | Paid work | θEM,age | 0.013 | 0.001 |
| Kenya | Paid work | θEM,marria | 0.033 | 0.026 |
| Kenya | Paid work | θEM,highes | 0.026 | 0.009 |
| Kenya | Paid work | θEM,births | -0.003 | 0.006 |
| Kenya | Paid work | θEM,urban | 0.166 | 0.032 |
| Kenya | Paid work | cov,CT3,EN | 0.014 | 0.003 |
| Kenya | Paid work | cov,CT2,EN | 0.012 | 0.003 |
| Kenya | Paid work | cov,CT1,EN | 0.022 | 0.004 |
| Kenya | Paid work | ε,EM3 | 0.174 | 0.004 |
| Kenya | Paid work | ε,EM2 | 0.17 | 0.004 |
| Kenya | Paid work | ε,CT1 | 0.21 | 0.004 |
| Kenya | Paid work | ε,EM3 | 0.185 | 0.004 |
| Kenya | Paid work | ε,EM2 | 0.206 | 0.004 |
| Kenya | Paid work | ε,EM1 | 0.221 | 0.003 |
| Kenya | Decision ar | vEM3 | 0.343 | 0.024 |
| Kenya | Decision ar | βEM,2,1 | 0.466 | 0.022 |
| Kenya | Decision ar | λEM,2,1 | -0.015 | 0.021 |
| Kenya | Decision ar | λEM,2,1 | 0.031 | 0.019 |
| Kenya | Decision ar | vEM2 | 0.375 | 0.023 |
| Kenya | Decision ar | βEM,1,0 | 0.476 | 0.021 |
| Kenya | Decision ar | λEM,1,0 | -0.017 | 0.019 |

| | | | | |
|---|---|---|---|---|
| Kenya | Decision ar | vCT1 | 0.766 | 0.064 |
| Kenya | Decision ar | θCT,age | -0.009 | 0.002 |
| Kenya | Decision ar | θCT,marria | 0.021 | 0.055 |
| Kenya | Decision ar | θCT,highes | 0.049 | 0.009 |
| Kenya | Decision ar | θCT,births | 0.02 | 0.007 |
| Kenya | Decision ar | θCT,urban | 0.09 | 0.025 |
| Kenya | Decision ar | vEM3 | 0.464 | 0.031 |
| Kenya | Decision ar | λEM,2,1 | 0.019 | 0.02 |
| Kenya | Decision ar | βEM,2,1 | 0.354 | 0.029 |
| Kenya | Decision ar | λEM,2,1 | -0.02 | 0.021 |
| Kenya | Decision ar | vEM2 | 0.548 | 0.027 |
| Kenya | Decision ar | λEM,1,0 | -0.03 | 0.022 |
| Kenya | Decision ar | βEM,1,0 | 0.289 | 0.024 |
| Kenya | Decision ar | vEM1 | 0.388 | 0.075 |
| Kenya | Decision ar | θEM,age | 0.005 | 0.002 |
| Kenya | Decision ar | θEM,marria | 0.128 | 0.064 |
| Kenya | Decision ar | θEM,highes | 0.011 | 0.009 |
| Kenya | Decision ar | θEM,births | 0.001 | 0.006 |
| Kenya | Decision ar | θEM,urban | 0.02 | 0.037 |
| Kenya | Decision ar | cov,CT3,EM | 0.008 | 0.004 |
| Kenya | Decision ar | cov,CT2,EM | 0.002 | 0.004 |
| Kenya | Decision ar | cov,CT1,EM | 0.004 | 0.005 |
| Kenya | Decision ar | ε,EM3 | 0.174 | 0.005 |
| Kenya | Decision ar | ε,EM2 | 0.167 | 0.005 |
| Kenya | Decision ar | ε,CT1 | 0.216 | 0.004 |
| Kenya | Decision ar | ε,EM3 | 0.176 | 0.008 |
| Kenya | Decision ar | ε,EM2 | 0.18 | 0.008 |
| Kenya | Decision ar | ε,EM1 | 0.209 | 0.006 |
| Kenya | Decision ar | vEM3 | 0.34 | 0.033 |
| Kenya | Decision ar | βEM,2,1 | 0.466 | 0.022 |
| Kenya | Decision ar | λEM,2,1 | -0.008 | 0.027 |
| Kenya | Decision ar | λEM,2,1 | 0.024 | 0.023 |
| Kenya | Decision ar | vEM2 | 0.363 | 0.024 |
| Kenya | Decision ar | βEM,1,0 | 0.477 | 0.021 |
| Kenya | Decision ar | λEM,1,0 | -0.001 | 0.02 |
| Kenya | Decision ar | vCT1 | 0.776 | 0.063 |
| Kenya | Decision ar | θCT,age | -0.009 | 0.002 |
| Kenya | Decision ar | θCT,marria | 0.017 | 0.054 |
| Kenya | Decision ar | θCT,highes | 0.049 | 0.009 |
| Kenya | Decision ar | θCT,births | 0.02 | 0.007 |
| Kenya | Decision ar | θCT,urban | 0.091 | 0.025 |
| Kenya | Decision ar | vEM3 | 0.555 | 0.032 |
| Kenya | Decision ar | λEM,2,1 | 0.031 | 0.019 |
| Kenya | Decision ar | βEM,2,1 | 0.327 | 0.033 |

| | | | | |
|---|---|---|---|---|
| Kenya | Decision a | λEM,2,1 | -0.015 | 0.017 |
| Kenya | Decision a | vEM2 | 0.677 | 0.027 |
| Kenya | Decision a | λEM,1,0 | -0.01 | 0.019 |
| Kenya | Decision a | βEM,1,0 | 0.213 | 0.026 |
| Kenya | Decision a | vEM1 | 0.451 | 0.061 |
| Kenya | Decision a | θEM,age | 0.008 | 0.001 |
| Kenya | Decision a | θEM,marria | 0.115 | 0.058 |
| Kenya | Decision a | θEM,highe | 0.006 | 0.007 |
| Kenya | Decision a | θEM,births | -0.007 | 0.005 |
| Kenya | Decision a | θEM,urban | 0.023 | 0.026 |
| Kenya | Decision a | cov,CT3,EN | 0.006 | 0.003 |
| Kenya | Decision a | cov,CT2,EN | 0.002 | 0.003 |
| Kenya | Decision a | cov,CT1,EN | 0 | 0.004 |
| Kenya | Decision a | ε,EM3 | 0.174 | 0.005 |
| Kenya | Decision a | ε,EM2 | 0.167 | 0.005 |
| Kenya | Decision a | ε,CT1 | 0.216 | 0.004 |
| Kenya | Decision a | ε,EM3 | 0.12 | 0.008 |
| Kenya | Decision a | ε,EM2 | 0.127 | 0.008 |
| Kenya | Decision a | ε,EM1 | 0.154 | 0.008 |
| Kenya | Decision a | vEM3 | 0.236 | 0.015 |
| Kenya | Decision a | βEM,2,1 | 0.515 | 0.019 |
| Kenya | Decision a | λEM,2,1 | 0.088 | 0.016 |
| Kenya | Decision a | λEM,2,1 | 0.022 | 0.02 |
| Kenya | Decision a | vEM2 | 0.259 | 0.013 |
| Kenya | Decision a | βEM,1,0 | 0.555 | 0.017 |
| Kenya | Decision a | λEM,1,0 | 0.052 | 0.017 |
| Kenya | Decision a | vCT1 | 0.132 | 0.046 |
| Kenya | Decision a | θCT,age | -0.002 | 0.002 |
| Kenya | Decision a | θCT,marria | 0.368 | 0.033 |
| Kenya | Decision a | θCT,highes | 0.045 | 0.009 |
| Kenya | Decision a | θCT,births | 0.031 | 0.007 |
| Kenya | Decision a | θCT,urban | 0.091 | 0.025 |
| Kenya | Decision a | vEM3 | 0.159 | 0.017 |
| Kenya | Decision a | λEM,2,1 | 0.131 | 0.02 |
| Kenya | Decision a | βEM,2,1 | 0.484 | 0.021 |
| Kenya | Decision a | λEM,2,1 | 0.042 | 0.021 |
| Kenya | Decision a | vEM2 | 0.232 | 0.017 |
| Kenya | Decision a | λEM,1,0 | 0.154 | 0.019 |
| Kenya | Decision a | βEM,1,0 | 0.432 | 0.021 |
| Kenya | Decision a | vEM1 | -0.175 | 0.043 |
| Kenya | Decision a | θEM,age | 0.008 | 0.002 |
| Kenya | Decision a | θEM,marria | 0.24 | 0.031 |
| Kenya | Decision a | θEM,highe | 0.032 | 0.009 |
| Kenya | Decision a | θEM,births | 0.004 | 0.007 |

| | | | | |
|---|---|---|---|---|
| Kenya | Decision a | θEM,urban | 0.116 | 0.034 |
| Kenya | Decision a | cov,CT3,EM | 0.012 | 0.004 |
| Kenya | Decision a | cov,CT2,EM | 0.017 | 0.003 |
| Kenya | Decision a | cov,CT1,EM | 0.02 | 0.005 |
| Kenya | Decision a | ε,EM3 | 0.169 | 0.005 |
| Kenya | Decision a | ε,EM2 | 0.163 | 0.005 |
| Kenya | Decision a | ε,CT1 | 0.209 | 0.005 |
| Kenya | Decision a | ε,EM3 | 0.176 | 0.005 |
| Kenya | Decision a | ε,EM2 | 0.195 | 0.004 |
| Kenya | Decision a | ε,EM1 | 0.196 | 0.005 |
| Kenya | Decision a | vEM3 | 0.338 | 0.022 |
| Kenya | Decision a | βEM,2,1 | 0.464 | 0.021 |
| Kenya | Decision a | λEM,2,1 | 0.021 | 0.022 |
| Kenya | Decision a | λEM,2,1 | 0.009 | 0.017 |
| Kenya | Decision a | vEM2 | 0.381 | 0.02 |
| Kenya | Decision a | βEM,1,0 | 0.474 | 0.021 |
| Kenya | Decision a | λEM,1,0 | -0.031 | 0.019 |
| Kenya | Decision a | vCT1 | 0.772 | 0.06 |
| Kenya | Decision a | θCT,age | -0.009 | 0.002 |
| Kenya | Decision a | θCT,marria | 0.011 | 0.05 |
| Kenya | Decision a | θCT,highes | 0.049 | 0.009 |
| Kenya | Decision a | θCT,births | 0.021 | 0.007 |
| Kenya | Decision a | θCT,urban | 0.092 | 0.025 |
| Kenya | Decision a | vEM3 | 0.333 | 0.027 |
| Kenya | Decision a | λEM,2,1 | 0.006 | 0.025 |
| Kenya | Decision a | βEM,2,1 | 0.339 | 0.026 |
| Kenya | Decision a | λEM,2,1 | 0.007 | 0.026 |
| Kenya | Decision a | vEM2 | 0.391 | 0.026 |
| Kenya | Decision a | λEM,1,0 | 0.007 | 0.022 |
| Kenya | Decision a | βEM,1,0 | 0.298 | 0.024 |
| Kenya | Decision a | vEM1 | 0.43 | 0.073 |
| Kenya | Decision a | θEM,age | 0.003 | 0.002 |
| Kenya | Decision a | θEM,marria | 0.041 | 0.05 |
| Kenya | Decision a | θEM,highes | 0 | 0.011 |
| Kenya | Decision a | θEM,births | -0.01 | 0.006 |
| Kenya | Decision a | θEM,urban | 0.037 | 0.04 |
| Kenya | Decision a | cov,CT3,EM | 0.003 | 0.005 |
| Kenya | Decision a | cov,CT2,EM | 0.01 | 0.004 |
| Kenya | Decision a | cov,CT1,EM | 0 | 0.005 |
| Kenya | Decision a | ε,EM3 | 0.174 | 0.005 |
| Kenya | Decision a | ε,EM2 | 0.167 | 0.005 |
| Kenya | Decision a | ε,CT1 | 0.216 | 0.004 |
| Kenya | Decision a | ε,EM3 | 0.221 | 0.004 |
| Kenya | Decision a | ε,EM2 | 0.225 | 0.004 |

| | | | | |
|---|---|---|---|---|
| Kenya | Decision ar | ε,EM1 | 0.248 | 0.001 |
| Kenya | Decision ar | vEM3 | 0.252 | 0.029 |
| Kenya | Decision ar | βEM,2,1 | 0.539 | 0.016 |
| Kenya | Decision ar | λEM,2,1 | 0.049 | 0.024 |
| Kenya | Decision ar | λEM,2,1 | -0.043 | 0.026 |
| Kenya | Decision ar | vEM2 | 0.255 | 0.02 |
| Kenya | Decision ar | βEM,1,0 | 0.554 | 0.015 |
| Kenya | Decision ar | λEM,1,0 | 0.014 | 0.02 |
| Kenya | Decision ar | vCT1 | 0.164 | 0.038 |
| Kenya | Decision ar | θCT,age | -0.001 | 0.001 |
| Kenya | Decision ar | θCT,marria | 0.335 | 0.022 |
| Kenya | Decision ar | θCT,highes | 0.038 | 0.008 |
| Kenya | Decision ar | θCT,births | 0.031 | 0.006 |
| Kenya | Decision ar | θCT,urban | 0.101 | 0.024 |
| Kenya | Decision ar | vEM3 | 0.597 | 0.043 |
| Kenya | Decision ar | λEM,2,1 | 0.007 | 0.012 |
| Kenya | Decision ar | βEM,2,1 | 0.33 | 0.048 |
| Kenya | Decision ar | λEM,2,1 | 0.002 | 0.011 |
| Kenya | Decision ar | vEM2 | 0.701 | 0.034 |
| Kenya | Decision ar | λEM,1,0 | 0.009 | 0.014 |
| Kenya | Decision ar | βEM,1,0 | 0.22 | 0.038 |
| Kenya | Decision ar | vEM1 | 0.851 | 0.026 |
| Kenya | Decision ar | θEM,age | 0.001 | 0.001 |
| Kenya | Decision ar | θEM,marria | -0.026 | 0.013 |
| Kenya | Decision ar | θEM,highes | 0.025 | 0.006 |
| Kenya | Decision ar | θEM,births | -0.011 | 0.005 |
| Kenya | Decision ar | θEM,urban | 0.021 | 0.022 |
| Kenya | Decision ar | cov,CT3,EM | 0.003 | 0.002 |
| Kenya | Decision ar | cov,CT2,EM | 0.005 | 0.002 |
| Kenya | Decision ar | cov,CT1,EM | 0.011 | 0.003 |
| Kenya | Decision ar | ε,EM3 | 0.175 | 0.004 |
| Kenya | Decision ar | ε,EM2 | 0.17 | 0.004 |
| Kenya | Decision ar | ε,CT1 | 0.21 | 0.004 |
| Kenya | Decision ar | ε,EM3 | 0.081 | 0.007 |
| Kenya | Decision ar | ε,EM2 | 0.085 | 0.009 |
| Kenya | Decision ar | ε,EM1 | 0.104 | 0.008 |
| Kenya | Decision ar | vEM3 | 0.207 | 0.025 |
| Kenya | Decision ar | βEM,2,1 | 0.535 | 0.017 |
| Kenya | Decision ar | λEM,2,1 | 0.047 | 0.022 |
| Kenya | Decision ar | λEM,2,1 | 0.013 | 0.019 |
| Kenya | Decision ar | vEM2 | 0.19 | 0.019 |
| Kenya | Decision ar | βEM,1,0 | 0.549 | 0.015 |
| Kenya | Decision ar | λEM,1,0 | 0.091 | 0.02 |
| Kenya | Decision ar | vCT1 | 0.166 | 0.039 |

| | | | | |
|---|---|---|---|---|
| Kenya | Decision ar | θCT,age | -0.001 | 0.002 |
| Kenya | Decision ar | θCT,marria | 0.334 | 0.022 |
| Kenya | Decision ar | θCT,highes | 0.038 | 0.008 |
| Kenya | Decision ar | θCT,births | 0.031 | 0.006 |
| Kenya | Decision ar | θCT,urban | 0.101 | 0.025 |
| Kenya | Decision ar | vEM3 | 0.68 | 0.037 |
| Kenya | Decision ar | λEM,2,1 | 0.042 | 0.01 |
| Kenya | Decision ar | βEM,2,1 | 0.234 | 0.039 |
| Kenya | Decision ar | λEM,2,1 | -0.007 | 0.011 |
| Kenya | Decision ar | vEM2 | 0.617 | 0.03 |
| Kenya | Decision ar | λEM,1,0 | 0.037 | 0.012 |
| Kenya | Decision ar | βEM,1,0 | 0.286 | 0.032 |
| Kenya | Decision ar | vEM1 | 0.789 | 0.027 |
| Kenya | Decision ar | θEM,age | 0.002 | 0.001 |
| Kenya | Decision ar | θEM,marria | -0.005 | 0.012 |
| Kenya | Decision ar | θEM,highes | 0.028 | 0.005 |
| Kenya | Decision ar | θEM,births | -0.001 | 0.004 |
| Kenya | Decision ar | θEM,urban | 0.036 | 0.015 |
| Kenya | Decision ar | cov,CT3,EM | 0.007 | 0.002 |
| Kenya | Decision ar | cov,CT2,EM | 0.006 | 0.002 |
| Kenya | Decision ar | cov,CT1,EM | 0.012 | 0.003 |
| Kenya | Decision ar | ε,EM3 | 0.175 | 0.004 |
| Kenya | Decision ar | ε,EM2 | 0.17 | 0.004 |
| Kenya | Decision ar | ε,CT1 | 0.211 | 0.004 |
| Kenya | Decision ar | ε,EM3 | 0.077 | 0.007 |
| Kenya | Decision ar | ε,EM2 | 0.089 | 0.007 |
| Kenya | Decision ar | ε,EM1 | 0.094 | 0.007 |
| Uganda | Paid work | vEM3 | 0.233 | 0.023 |
| Uganda | Paid work | βEM,2,1 | 0.442 | 0.025 |
| Uganda | Paid work | λEM,2,1 | 0.047 | 0.026 |
| Uganda | Paid work | λEM,2,1 | -0.023 | 0.028 |
| Uganda | Paid work | vEM2 | 0.282 | 0.028 |
| Uganda | Paid work | βEM,1,0 | 0.415 | 0.032 |
| Uganda | Paid work | λEM,1,0 | -0.033 | 0.035 |
| Uganda | Paid work | vCT1 | 0.37 | 0.056 |
| Uganda | Paid work | θCT,age | -0.007 | 0.002 |
| Uganda | Paid work | θCT,marria | 0.044 | 0.043 |
| Uganda | Paid work | θCT,highes | 0.051 | 0.024 |
| Uganda | Paid work | θCT,births | 0.046 | 0.01 |
| Uganda | Paid work | θCT,urban | 0.09 | 0.039 |
| Uganda | Paid work | vEM3 | 0.35 | 0.029 |
| Uganda | Paid work | λEM,2,1 | 0.004 | 0.035 |
| Uganda | Paid work | βEM,2,1 | 0.452 | 0.029 |
| Uganda | Paid work | λEM,2,1 | 0.018 | 0.035 |

| | | | | |
|---|---|---|---|---|
| Uganda | Paid work | vEM2 | 0.351 | 0.032 |
| Uganda | Paid work | λEM,1,0 | 0.023 | 0.03 |
| Uganda | Paid work | βEM,1,0 | 0.366 | 0.039 |
| Uganda | Paid work | vEM1 | 0.091 | 0.072 |
| Uganda | Paid work | θEM,age | 0.014 | 0.003 |
| Uganda | Paid work | θEM,marria | -0.039 | 0.035 |
| Uganda | Paid work | θEM,highes | 0.03 | 0.02 |
| Uganda | Paid work | θEM,births | -0.028 | 0.011 |
| Uganda | Paid work | θEM,urban | -0.025 | 0.074 |
| Uganda | Paid work | cov,CT3,EN | -0.002 | 0.005 |
| Uganda | Paid work | cov,CT2,EN | 0.005 | 0.006 |
| Uganda | Paid work | cov,CT1,EN | 0.014 | 0.007 |
| Uganda | Paid work | ε,EM3 | 0.198 | 0.005 |
| Uganda | Paid work | ε,EM2 | 0.205 | 0.007 |
| Uganda | Paid work | ε,CT1 | 0.237 | 0.004 |
| Uganda | Paid work | ε,EM3 | 0.189 | 0.007 |
| Uganda | Paid work | ε,EM2 | 0.216 | 0.007 |
| Uganda | Paid work | ε,EM1 | 0.239 | 0.004 |
| Uganda | Decision ar | vEM3 | 0.356 | 0.033 |
| Uganda | Decision ar | βEM,2,1 | 0.398 | 0.029 |
| Uganda | Decision ar | λEM,2,1 | -0.019 | 0.031 |
| Uganda | Decision ar | λEM,2,1 | -0.054 | 0.034 |
| Uganda | Decision ar | vEM2 | 0.397 | 0.038 |
| Uganda | Decision ar | βEM,1,0 | 0.344 | 0.04 |
| Uganda | Decision ar | λEM,1,0 | -0.078 | 0.032 |
| Uganda | Decision ar | vCT1 | 0.517 | 0.089 |
| Uganda | Decision ar | θCT,age | -0.005 | 0.004 |
| Uganda | Decision ar | θCT,marria | -0.046 | 0.039 |
| Uganda | Decision ar | θCT,highes | 0.061 | 0.028 |
| Uganda | Decision ar | θCT,births | 0.025 | 0.013 |
| Uganda | Decision ar | θCT,urban | 0.109 | 0.04 |
| Uganda | Decision ar | vEM3 | 0.315 | 0.042 |
| Uganda | Decision ar | λEM,2,1 | -0.077 | 0.038 |
| Uganda | Decision ar | βEM,2,1 | 0.44 | 0.039 |
| Uganda | Decision ar | λEM,2,1 | 0.053 | 0.041 |
| Uganda | Decision ar | vEM2 | 0.305 | 0.029 |
| Uganda | Decision ar | λEM,1,0 | -0.01 | 0.031 |
| Uganda | Decision ar | βEM,1,0 | 0.458 | 0.033 |
| Uganda | Decision ar | vEM1 | 0.137 | 0.094 |
| Uganda | Decision ar | θEM,age | 0.014 | 0.004 |
| Uganda | Decision ar | θEM,marria | 0.038 | 0.05 |
| Uganda | Decision ar | θEM,highes | -0.018 | 0.018 |
| Uganda | Decision ar | θEM,births | -0.012 | 0.01 |
| Uganda | Decision ar | θEM,urban | 0.076 | 0.071 |

| | | | | |
|---|---|---|---|---|
| Uganda | Decision a | cov,CT3,EN | 0.002 | 0.007 |
| Uganda | Decision a | cov,CT2,EN | -0.006 | 0.008 |
| Uganda | Decision a | cov,CT1,EN | -0.008 | 0.012 |
| Uganda | Decision a | ε,EM3 | 0.207 | 0.006 |
| Uganda | Decision a | ε,EM2 | 0.217 | 0.007 |
| Uganda | Decision a | ε,CT1 | 0.242 | 0.004 |
| Uganda | Decision a | ε,EM3 | 0.198 | 0.009 |
| Uganda | Decision a | ε,EM2 | 0.196 | 0.008 |
| Uganda | Decision a | ε,EM1 | 0.239 | 0.004 |
| Uganda | Decision a | vEM3 | 0.33 | 0.045 |
| Uganda | Decision a | βEM,2,1 | 0.41 | 0.029 |
| Uganda | Decision a | λEM,2,1 | -0.01 | 0.04 |
| Uganda | Decision a | λEM,2,1 | -0.022 | 0.039 |
| Uganda | Decision a | vEM2 | 0.388 | 0.042 |
| Uganda | Decision a | βEM,1,0 | 0.35 | 0.041 |
| Uganda | Decision a | λEM,1,0 | -0.048 | 0.041 |
| Uganda | Decision a | vCT1 | 0.507 | 0.087 |
| Uganda | Decision a | θCT,age | -0.005 | 0.004 |
| Uganda | Decision a | θCT,marria | -0.047 | 0.039 |
| Uganda | Decision a | θCT,highes | 0.062 | 0.028 |
| Uganda | Decision a | θCT,births | 0.026 | 0.013 |
| Uganda | Decision a | θCT,urban | 0.106 | 0.04 |
| Uganda | Decision a | vEM3 | 0.525 | 0.039 |
| Uganda | Decision a | λEM,2,1 | -0.04 | 0.038 |
| Uganda | Decision a | βEM,2,1 | 0.308 | 0.05 |
| Uganda | Decision a | λEM,2,1 | 0.024 | 0.051 |
| Uganda | Decision a | vEM2 | 0.537 | 0.053 |
| Uganda | Decision a | λEM,1,0 | -0.004 | 0.035 |
| Uganda | Decision a | βEM,1,0 | 0.274 | 0.054 |
| Uganda | Decision a | vEM1 | 0.313 | 0.091 |
| Uganda | Decision a | θEM,age | 0.012 | 0.004 |
| Uganda | Decision a | θEM,marria | 0.035 | 0.036 |
| Uganda | Decision a | θEM,highes | 0.026 | 0.016 |
| Uganda | Decision a | θEM,births | -0.011 | 0.012 |
| Uganda | Decision a | θEM,urban | 0.013 | 0.059 |
| Uganda | Decision a | cov,CT3,EN | -0.015 | 0.007 |
| Uganda | Decision a | cov,CT2,EN | 0.005 | 0.007 |
| Uganda | Decision a | cov,CT1,EN | 0.003 | 0.011 |
| Uganda | Decision a | ε,EM3 | 0.207 | 0.006 |
| Uganda | Decision a | ε,EM2 | 0.217 | 0.007 |
| Uganda | Decision a | ε,CT1 | 0.242 | 0.004 |
| Uganda | Decision a | ε,EM3 | 0.173 | 0.012 |
| Uganda | Decision a | ε,EM2 | 0.183 | 0.01 |
| Uganda | Decision a | ε,EM1 | 0.203 | 0.01 |

| | | | | |
|---|---|---|---|---|
| Uganda | Decision a | vEM3 | 0.237 | 0.044 |
| Uganda | Decision a | βEM,2,1 | 0.435 | 0.025 |
| Uganda | Decision a | λEM,2,1 | 0.048 | 0.038 |
| Uganda | Decision a | λEM,2,1 | -0.029 | 0.034 |
| Uganda | Decision a | vEM2 | 0.31 | 0.04 |
| Uganda | Decision a | βEM,1,0 | 0.408 | 0.034 |
| Uganda | Decision a | λEM,1,0 | -0.049 | 0.037 |
| Uganda | Decision a | vCT1 | 0.39 | 0.055 |
| Uganda | Decision a | θCT,age | -0.007 | 0.002 |
| Uganda | Decision a | θCT,marria | 0.043 | 0.045 |
| Uganda | Decision a | θCT,highes | 0.05 | 0.025 |
| Uganda | Decision a | θCT,births | 0.045 | 0.01 |
| Uganda | Decision a | θCT,urban | 0.089 | 0.041 |
| Uganda | Decision a | vEM3 | 0.64 | 0.035 |
| Uganda | Decision a | λEM,2,1 | 0.009 | 0.02 |
| Uganda | Decision a | βEM,2,1 | 0.213 | 0.036 |
| Uganda | Decision a | λEM,2,1 | -0.014 | 0.025 |
| Uganda | Decision a | vEM2 | 0.68 | 0.036 |
| Uganda | Decision a | λEM,1,0 | 0.01 | 0.024 |
| Uganda | Decision a | βEM,1,0 | 0.158 | 0.037 |
| Uganda | Decision a | vEM1 | 0.742 | 0.062 |
| Uganda | Decision a | θEM,age | -0.001 | 0.002 |
| Uganda | Decision a | θEM,marria | 0.062 | 0.036 |
| Uganda | Decision a | θEM,highes | 0.054 | 0.018 |
| Uganda | Decision a | θEM,births | -0.018 | 0.012 |
| Uganda | Decision a | θEM,urban | 0.015 | 0.051 |
| Uganda | Decision a | cov,CT3,EM | 0.012 | 0.005 |
| Uganda | Decision a | cov,CT2,EM | 0.015 | 0.008 |
| Uganda | Decision a | cov,CT1,EM | 0.001 | 0.006 |
| Uganda | Decision a | ε,EM3 | 0.2 | 0.005 |
| Uganda | Decision a | ε,EM2 | 0.207 | 0.007 |
| Uganda | Decision a | ε,CT1 | 0.238 | 0.004 |
| Uganda | Decision a | ε,EM3 | 0.147 | 0.011 |
| Uganda | Decision a | ε,EM2 | 0.154 | 0.011 |
| Uganda | Decision a | ε,EM1 | 0.183 | 0.011 |
| Uganda | Decision a | vEM3 | 0.158 | 0.04 |
| Uganda | Decision a | βEM,2,1 | 0.431 | 0.024 |
| Uganda | Decision a | λEM,2,1 | 0.069 | 0.036 |
| Uganda | Decision a | λEM,2,1 | 0.047 | 0.032 |
| Uganda | Decision a | vEM2 | 0.25 | 0.037 |
| Uganda | Decision a | βEM,1,0 | 0.4 | 0.034 |
| Uganda | Decision a | λEM,1,0 | 0.037 | 0.038 |
| Uganda | Decision a | vCT1 | 0.408 | 0.057 |
| Uganda | Decision a | θCT,age | -0.007 | 0.003 |

| | | | | |
|---|---|---|---|---|
| Uganda | Decision ai | θCT,marria | 0.043 | 0.045 |
| Uganda | Decision ai | θCT,highes | 0.05 | 0.025 |
| Uganda | Decision ai | θCT,births | 0.045 | 0.01 |
| Uganda | Decision ai | θCT,urban | 0.087 | 0.041 |
| Uganda | Decision ai | vEM3 | 0.618 | 0.036 |
| Uganda | Decision ai | λEM,2,1 | 0.004 | 0.022 |
| Uganda | Decision ai | βEM,2,1 | 0.285 | 0.037 |
| Uganda | Decision ai | λEM,2,1 | -0.023 | 0.023 |
| Uganda | Decision ai | vEM2 | 0.643 | 0.041 |
| Uganda | Decision ai | λEM,1,0 | 0.046 | 0.019 |
| Uganda | Decision ai | βEM,1,0 | 0.201 | 0.042 |
| Uganda | Decision ai | vEM1 | 0.829 | 0.059 |
| Uganda | Decision ai | θEM,age | -0.001 | 0.002 |
| Uganda | Decision ai | θEM,marria | 0.078 | 0.025 |
| Uganda | Decision ai | θEM,highes | 0.016 | 0.011 |
| Uganda | Decision ai | θEM,births | -0.01 | 0.009 |
| Uganda | Decision ai | θEM,urban | 0.053 | 0.032 |
| Uganda | Decision ai | cov,CT3,EN | 0.011 | 0.006 |
| Uganda | Decision ai | cov,CT2,EN | 0.009 | 0.006 |
| Uganda | Decision ai | cov,CT1,EN | 0.007 | 0.006 |
| Uganda | Decision ai | ε,EM3 | 0.199 | 0.005 |
| Uganda | Decision ai | ε,EM2 | 0.209 | 0.007 |
| Uganda | Decision ai | ε,CT1 | 0.239 | 0.004 |
| Uganda | Decision ai | ε,EM3 | 0.119 | 0.011 |
| Uganda | Decision ai | ε,EM2 | 0.134 | 0.01 |
| Uganda | Decision ai | ε,EM1 | 0.135 | 0.009 |
| Burkina | Paid work | vEM3 | 0.163 | 0.018 |
| Burkina | Paid work | βEM,2,1 | 0.466 | 0.023 |
| Burkina | Paid work | λEM,2,1 | 0.015 | 0.021 |
| Burkina | Paid work | λEM,2,1 | 0 | 0.026 |
| Burkina | Paid work | vEM2 | 0.184 | 0.018 |
| Burkina | Paid work | βEM,1,0 | 0.469 | 0.02 |
| Burkina | Paid work | λEM,1,0 | 0.023 | 0.024 |
| Burkina | Paid work | vCT1 | 0.245 | 0.052 |
| Burkina | Paid work | θCT,age | -0.005 | 0.002 |
| Burkina | Paid work | θCT,marria | 0.088 | 0.045 |
| Burkina | Paid work | θCT,highes | 0.019 | 0.007 |
| Burkina | Paid work | θCT,births | 0.037 | 0.009 |
| Burkina | Paid work | θCT,urban | 0.165 | 0.034 |
| Burkina | Paid work | vEM3 | 0.243 | 0.03 |
| Burkina | Paid work | λEM,2,1 | 0.035 | 0.02 |
| Burkina | Paid work | βEM,2,1 | 0.489 | 0.036 |
| Burkina | Paid work | λEM,2,1 | 0.02 | 0.029 |
| Burkina | Paid work | vEM2 | 0.292 | 0.035 |

| | | | | |
|---|---|---|---|---|
| Burkina | Paid work | λEM,1,0 | 0.035 | 0.03 |
| Burkina | Paid work | βEM,1,0 | 0.422 | 0.046 |
| Burkina | Paid work | vEM1 | -0.07 | 0.058 |
| Burkina | Paid work | θEM,age | 0.015 | 0.003 |
| Burkina | Paid work | θEM,marria | 0.075 | 0.047 |
| Burkina | Paid work | θEM,highes | -0.004 | 0.007 |
| Burkina | Paid work | θEM,births | -0.034 | 0.01 |
| Burkina | Paid work | θEM,urban | 0.182 | 0.046 |
| Burkina | Paid work | cov,CT3,EN | 0.002 | 0.005 |
| Burkina | Paid work | cov,CT2,EN | 0.009 | 0.006 |
| Burkina | Paid work | cov,CT1,EN | 0.012 | 0.006 |
| Burkina | Paid work | ε,EM3 | 0.172 | 0.007 |
| Burkina | Paid work | ε,EM2 | 0.178 | 0.006 |
| Burkina | Paid work | ε,CT1 | 0.214 | 0.009 |
| Burkina | Paid work | ε,EM3 | 0.189 | 0.009 |
| Burkina | Paid work | ε,EM2 | 0.206 | 0.009 |
| Burkina | Paid work | ε,EM1 | 0.215 | 0.012 |
| Burkina | Decision ar | vEM3 | 0.206 | 0.022 |
| Burkina | Decision ar | βEM,2,1 | 0.437 | 0.026 |
| Burkina | Decision ar | λEM,2,1 | 0.014 | 0.029 |
| Burkina | Decision ar | λEM,2,1 | -0.011 | 0.033 |
| Burkina | Decision ar | vEM2 | 0.204 | 0.023 |
| Burkina | Decision ar | βEM,1,0 | 0.439 | 0.024 |
| Burkina | Decision ar | λEM,1,0 | 0.046 | 0.03 |
| Burkina | Decision ar | vCT1 | 0.536 | 0.086 |
| Burkina | Decision ar | θCT,age | -0.007 | 0.002 |
| Burkina | Decision ar | θCT,marria | -0.098 | 0.074 |
| Burkina | Decision ar | θCT,highes | 0.015 | 0.006 |
| Burkina | Decision ar | θCT,births | 0.031 | 0.009 |
| Burkina | Decision ar | θCT,urban | 0.184 | 0.04 |
| Burkina | Decision ar | vEM3 | 0.225 | 0.035 |
| Burkina | Decision ar | λEM,2,1 | 0.048 | 0.029 |
| Burkina | Decision ar | βEM,2,1 | 0.334 | 0.038 |
| Burkina | Decision ar | λEM,2,1 | 0.017 | 0.035 |
| Burkina | Decision ar | vEM2 | 0.3 | 0.033 |
| Burkina | Decision ar | λEM,1,0 | 0.043 | 0.041 |
| Burkina | Decision ar | βEM,1,0 | 0.242 | 0.039 |
| Burkina | Decision ar | vEM1 | -0.08 | 0.1 |
| Burkina | Decision ar | θEM,age | 0.009 | 0.003 |
| Burkina | Decision ar | θEM,marria | 0.125 | 0.096 |
| Burkina | Decision ar | θEM,highes | 0.017 | 0.005 |
| Burkina | Decision ar | θEM,births | 0.008 | 0.011 |
| Burkina | Decision ar | θEM,urban | 0.024 | 0.048 |
| Burkina | Decision ar | cov,CT3,EN | 0.004 | 0.008 |

| | | | | |
|---|---|---|---|---|
| Burkina | Decision ar | cov,CT2,EN | 0.004 | 0.006 |
| Burkina | Decision ar | cov,CT1,EN | 0.003 | 0.008 |
| Burkina | Decision ar | ε,EM3 | 0.19 | 0.008 |
| Burkina | Decision ar | ε,EM2 | 0.192 | 0.007 |
| Burkina | Decision ar | ε,CT1 | 0.231 | 0.007 |
| Burkina | Decision ar | ε,EM3 | 0.209 | 0.008 |
| Burkina | Decision ar | ε,EM2 | 0.227 | 0.007 |
| Burkina | Decision ar | ε,EM1 | 0.222 | 0.01 |
| Burkina | Decision ar | vEM3 | 0.235 | 0.036 |
| Burkina | Decision ar | βEM,2,1 | 0.444 | 0.024 |
| Burkina | Decision ar | λEM,2,1 | -0.055 | 0.03 |
| Burkina | Decision ar | λEM,2,1 | 0.003 | 0.038 |
| Burkina | Decision ar | vEM2 | 0.2 | 0.028 |
| Burkina | Decision ar | βEM,1,0 | 0.445 | 0.022 |
| Burkina | Decision ar | λEM,1,0 | 0.02 | 0.026 |
| Burkina | Decision ar | vCT1 | 0.509 | 0.088 |
| Burkina | Decision ar | θCT,age | -0.007 | 0.002 |
| Burkina | Decision ar | θCT,marria | -0.103 | 0.077 |
| Burkina | Decision ar | θCT,highes | 0.016 | 0.006 |
| Burkina | Decision ar | θCT,births | 0.035 | 0.01 |
| Burkina | Decision ar | θCT,urban | 0.196 | 0.042 |
| Burkina | Decision ar | vEM3 | 0.494 | 0.04 |
| Burkina | Decision ar | λEM,2,1 | 0.012 | 0.02 |
| Burkina | Decision ar | βEM,2,1 | 0.341 | 0.041 |
| Burkina | Decision ar | λEM,2,1 | 0.038 | 0.028 |
| Burkina | Decision ar | vEM2 | 0.502 | 0.04 |
| Burkina | Decision ar | λEM,1,0 | -0.015 | 0.028 |
| Burkina | Decision ar | βEM,1,0 | 0.32 | 0.039 |
| Burkina | Decision ar | vEM1 | 0.647 | 0.075 |
| Burkina | Decision ar | θEM,age | 0.005 | 0.002 |
| Burkina | Decision ar | θEM,marria | -0.07 | 0.051 |
| Burkina | Decision ar | θEM,highes | 0 | 0.004 |
| Burkina | Decision ar | θEM,births | 0.006 | 0.009 |
| Burkina | Decision ar | θEM,urban | -0.04 | 0.039 |
| Burkina | Decision ar | cov,CT3,EN | -0.003 | 0.005 |
| Burkina | Decision ar | cov,CT2,EN | 0.009 | 0.005 |
| Burkina | Decision ar | cov,CT1,EN | -0.004 | 0.007 |
| Burkina | Decision ar | ε,EM3 | 0.185 | 0.008 |
| Burkina | Decision ar | ε,EM2 | 0.19 | 0.006 |
| Burkina | Decision ar | ε,CT1 | 0.226 | 0.009 |
| Burkina | Decision ar | ε,EM3 | 0.157 | 0.011 |
| Burkina | Decision ar | ε,EM2 | 0.173 | 0.01 |
| Burkina | Decision ar | ε,EM1 | 0.176 | 0.015 |
| Burkina | Decision ar | vEM3 | 0.168 | 0.018 |

| | | | | |
|---|---|---|---|---|
| Burkina | Decision a | βEM,2,1 | 0.455 | 0.026 |
| Burkina | Decision a | λEM,2,1 | 0.017 | 0.027 |
| Burkina | Decision a | λEM,2,1 | 0.007 | 0.026 |
| Burkina | Decision a | vEM2 | 0.198 | 0.019 |
| Burkina | Decision a | βEM,1,0 | 0.466 | 0.021 |
| Burkina | Decision a | λEM,1,0 | -0.004 | 0.029 |
| Burkina | Decision a | vCT1 | 0.306 | 0.061 |
| Burkina | Decision a | θCT,age | -0.005 | 0.002 |
| Burkina | Decision a | θCT,marria | 0.043 | 0.054 |
| Burkina | Decision a | θCT,highes | 0.015 | 0.007 |
| Burkina | Decision a | θCT,births | 0.036 | 0.01 |
| Burkina | Decision a | θCT,urban | 0.161 | 0.039 |
| Burkina | Decision a | vEM3 | 0.195 | 0.028 |
| Burkina | Decision a | λEM,2,1 | 0.053 | 0.027 |
| Burkina | Decision a | βEM,2,1 | 0.507 | 0.036 |
| Burkina | Decision a | λEM,2,1 | 0.014 | 0.03 |
| Burkina | Decision a | vEM2 | 0.223 | 0.034 |
| Burkina | Decision a | λEM,1,0 | 0.043 | 0.032 |
| Burkina | Decision a | βEM,1,0 | 0.429 | 0.053 |
| Burkina | Decision a | vEM1 | -0.153 | 0.059 |
| Burkina | Decision a | θEM,age | 0.012 | 0.003 |
| Burkina | Decision a | θEM,marria | 0.154 | 0.053 |
| Burkina | Decision a | θEM,highes | -0.009 | 0.007 |
| Burkina | Decision a | θEM,births | -0.023 | 0.01 |
| Burkina | Decision a | θEM,urban | 0.195 | 0.044 |
| Burkina | Decision a | cov,CT3,EM | 0.003 | 0.005 |
| Burkina | Decision a | cov,CT2,EM | 0.013 | 0.005 |
| Burkina | Decision a | cov,CT1,EM | 0.013 | 0.006 |
| Burkina | Decision a | ε,EM3 | 0.176 | 0.008 |
| Burkina | Decision a | ε,EM2 | 0.181 | 0.007 |
| Burkina | Decision a | ε,CT1 | 0.22 | 0.009 |
| Burkina | Decision a | ε,EM3 | 0.179 | 0.01 |
| Burkina | Decision a | ε,EM2 | 0.192 | 0.011 |
| Burkina | Decision a | ε,EM1 | 0.188 | 0.015 |
| Burkina | Decision a | vEM3 | 0.186 | 0.019 |
| Burkina | Decision a | βEM,2,1 | 0.438 | 0.024 |
| Burkina | Decision a | λEM,2,1 | 0.001 | 0.029 |
| Burkina | Decision a | λEM,2,1 | 0.039 | 0.036 |
| Burkina | Decision a | vEM2 | 0.21 | 0.02 |
| Burkina | Decision a | βEM,1,0 | 0.455 | 0.023 |
| Burkina | Decision a | λEM,1,0 | 0.007 | 0.029 |
| Burkina | Decision a | vCT1 | 0.51 | 0.085 |
| Burkina | Decision a | θCT,age | -0.007 | 0.002 |
| Burkina | Decision a | θCT,marria | -0.106 | 0.075 |

| | | | | |
|---|---|---|---|---|
| Burkina | Decision a | θCT,highes | 0.018 | 0.006 |
| Burkina | Decision a | θCT,births | 0.037 | 0.011 |
| Burkina | Decision a | θCT,urban | 0.195 | 0.042 |
| Burkina | Decision a | νEM3 | 0.125 | 0.025 |
| Burkina | Decision a | λEM,2,1 | -0.023 | 0.03 |
| Burkina | Decision a | βEM,2,1 | 0.306 | 0.045 |
| Burkina | Decision a | λEM,2,1 | 0.056 | 0.027 |
| Burkina | Decision a | νEM2 | 0.166 | 0.026 |
| Burkina | Decision a | λEM,1,0 | -0.011 | 0.026 |
| Burkina | Decision a | βEM,1,0 | 0.229 | 0.039 |
| Burkina | Decision a | νEM1 | 0.332 | 0.109 |
| Burkina | Decision a | θEM,age | 0.001 | 0.003 |
| Burkina | Decision a | θEM,marria | -0.097 | 0.117 |
| Burkina | Decision a | θEM,highes | 0.008 | 0.005 |
| Burkina | Decision a | θEM,births | 0.001 | 0.01 |
| Burkina | Decision a | θEM,urban | -0.106 | 0.043 |
| Burkina | Decision a | cov,CT3,EM | -0.002 | 0.004 |
| Burkina | Decision a | cov,CT2,EM | 0.004 | 0.005 |
| Burkina | Decision a | cov,CT1,EM | 0.018 | 0.011 |
| Burkina | Decision a | ε,EM3 | 0.186 | 0.008 |
| Burkina | Decision a | ε,EM2 | 0.187 | 0.007 |
| Burkina | Decision a | ε,CT1 | 0.225 | 0.009 |
| Burkina | Decision a | ε,EM3 | 0.147 | 0.013 |
| Burkina | Decision a | ε,EM2 | 0.165 | 0.012 |
| Burkina | Decision a | ε,EM1 | 0.199 | 0.013 |
| Burkina | Decision a | νEM3 | 0.138 | 0.023 |
| Burkina | Decision a | βEM,2,1 | 0.46 | 0.022 |
| Burkina | Decision a | λEM,2,1 | 0.044 | 0.021 |
| Burkina | Decision a | λEM,2,1 | 0.01 | 0.024 |
| Burkina | Decision a | νEM2 | 0.179 | 0.022 |
| Burkina | Decision a | βEM,1,0 | 0.469 | 0.02 |
| Burkina | Decision a | λEM,1,0 | 0.022 | 0.024 |
| Burkina | Decision a | νCT1 | 0.258 | 0.053 |
| Burkina | Decision a | θCT,age | -0.005 | 0.002 |
| Burkina | Decision a | θCT,marria | 0.08 | 0.046 |
| Burkina | Decision a | θCT,highes | 0.019 | 0.007 |
| Burkina | Decision a | θCT,births | 0.035 | 0.009 |
| Burkina | Decision a | θCT,urban | 0.163 | 0.034 |
| Burkina | Decision a | νEM3 | 0.362 | 0.046 |
| Burkina | Decision a | λEM,2,1 | 0.062 | 0.036 |
| Burkina | Decision a | βEM,2,1 | 0.35 | 0.05 |
| Burkina | Decision a | λEM,2,1 | 0.065 | 0.022 |
| Burkina | Decision a | νEM2 | 0.495 | 0.049 |
| Burkina | Decision a | λEM,1,0 | 0.058 | 0.033 |

| | | | | |
|---|---|---|---|---|
| Burkina | Decision a | βEM,1,0 | 0.26 | 0.055 |
| Burkina | Decision a | vEM1 | 0.756 | 0.068 |
| Burkina | Decision a | θEM,age | -0.006 | 0.002 |
| Burkina | Decision a | θEM,marria | -0.012 | 0.049 |
| Burkina | Decision a | θEM,highes | 0.011 | 0.006 |
| Burkina | Decision a | θEM,births | 0.005 | 0.012 |
| Burkina | Decision a | θEM,urban | 0.029 | 0.055 |
| Burkina | Decision a | cov,CT3,EM | 0.006 | 0.004 |
| Burkina | Decision a | cov,CT2,EM | 0.003 | 0.006 |
| Burkina | Decision a | cov,CT1,EM | 0.015 | 0.007 |
| Burkina | Decision a | ε,EM3 | 0.174 | 0.008 |
| Burkina | Decision a | ε,EM2 | 0.178 | 0.006 |
| Burkina | Decision a | ε,CT1 | 0.216 | 0.009 |
| Burkina | Decision a | ε,EM3 | 0.199 | 0.013 |
| Burkina | Decision a | ε,EM2 | 0.203 | 0.011 |
| Burkina | Decision a | ε,EM1 | 0.236 | 0.008 |
| Burkina | Decision a | vEM3 | 0.109 | 0.024 |
| Burkina | Decision a | βEM,2,1 | 0.461 | 0.024 |
| Burkina | Decision a | λEM,2,1 | 0.065 | 0.028 |
| Burkina | Decision a | λEM,2,1 | 0.018 | 0.021 |
| Burkina | Decision a | vEM2 | 0.14 | 0.023 |
| Burkina | Decision a | βEM,1,0 | 0.463 | 0.021 |
| Burkina | Decision a | λEM,1,0 | 0.075 | 0.025 |
| Burkina | Decision a | vCT1 | 0.268 | 0.054 |
| Burkina | Decision a | θCT,age | -0.005 | 0.002 |
| Burkina | Decision a | θCT,marria | 0.079 | 0.048 |
| Burkina | Decision a | θCT,highes | 0.019 | 0.007 |
| Burkina | Decision a | θCT,births | 0.036 | 0.01 |
| Burkina | Decision a | θCT,urban | 0.166 | 0.035 |
| Burkina | Decision a | vEM3 | 0.506 | 0.065 |
| Burkina | Decision a | λEM,2,1 | 0.026 | 0.021 |
| Burkina | Decision a | βEM,2,1 | 0.359 | 0.074 |
| Burkina | Decision a | λEM,2,1 | 0.037 | 0.019 |
| Burkina | Decision a | vEM2 | 0.525 | 0.051 |
| Burkina | Decision a | λEM,1,0 | 0.07 | 0.023 |
| Burkina | Decision a | βEM,1,0 | 0.316 | 0.056 |
| Burkina | Decision a | vEM1 | 0.714 | 0.055 |
| Burkina | Decision a | θEM,age | 0 | 0.002 |
| Burkina | Decision a | θEM,marria | 0.024 | 0.03 |
| Burkina | Decision a | θEM,highes | 0.01 | 0.005 |
| Burkina | Decision a | θEM,births | 0.002 | 0.007 |
| Burkina | Decision a | θEM,urban | 0.002 | 0.044 |
| Burkina | Decision a | cov,CT3,EM | 0.004 | 0.004 |
| Burkina | Decision a | cov,CT2,EM | 0.001 | 0.004 |

| | | | | |
|---|---|---|---|---|
| Burkina | Decision ai | cov,CT1,EN | 0.013 | 0.006 |
| Burkina | Decision ai | ε,EM3 | 0.173 | 0.007 |
| Burkina | Decision ai | ε,EM2 | 0.179 | 0.006 |
| Burkina | Decision ai | ε,CT1 | 0.217 | 0.009 |
| Burkina | Decision ai | ε,EM3 | 0.13 | 0.013 |
| Burkina | Decision ai | ε,EM2 | 0.146 | 0.011 |
| Burkina | Decision ai | ε,EM1 | 0.182 | 0.015 |
| DRC | Paid work | vEM3 | 0.222 | 0.021 |
| DRC | Paid work | βEM,2,1 | 0.443 | 0.025 |
| DRC | Paid work | λEM,2,1 | -0.024 | 0.029 |
| DRC | Paid work | λEM,2,1 | 0.01 | 0.033 |
| DRC | Paid work | vEM2 | 0.231 | 0.024 |
| DRC | Paid work | βEM,1,0 | 0.514 | 0.025 |
| DRC | Paid work | λEM,1,0 | 0.016 | 0.028 |
| DRC | Paid work | vCT1 | 0.421 | 0.065 |
| DRC | Paid work | θCT,age | -0.004 | 0.003 |
| DRC | Paid work | θCT,marria | 0.082 | 0.041 |
| DRC | Paid work | θCT,highes | 0.003 | 0.002 |
| DRC | Paid work | θCT,births | 0.039 | 0.01 |
| DRC | Paid work | vEM3 | 0.233 | 0.027 |
| DRC | Paid work | λEM,2,1 | 0.018 | 0.032 |
| DRC | Paid work | βEM,2,1 | 0.524 | 0.039 |
| DRC | Paid work | λEM,2,1 | 0.024 | 0.03 |
| DRC | Paid work | vEM2 | 0.289 | 0.037 |
| DRC | Paid work | λEM,1,0 | 0.04 | 0.035 |
| DRC | Paid work | βEM,1,0 | 0.481 | 0.031 |
| DRC | Paid work | vEM1 | -0.115 | 0.058 |
| DRC | Paid work | θEM,age | 0.02 | 0.002 |
| DRC | Paid work | θEM,marria | 0.068 | 0.048 |
| DRC | Paid work | θEM,highes | 0 | 0.005 |
| DRC | Paid work | θEM,births | -0.049 | 0.01 |
| DRC | Paid work | cov,CT3,EN | -0.001 | 0.005 |
| DRC | Paid work | cov,CT2,EN | 0.004 | 0.008 |
| DRC | Paid work | cov,CT1,EN | 0.022 | 0.008 |
| DRC | Paid work | ε,EM3 | 0.194 | 0.005 |
| DRC | Paid work | ε,EM2 | 0.183 | 0.006 |
| DRC | Paid work | ε,CT1 | 0.237 | 0.004 |
| DRC | Paid work | ε,EM3 | 0.18 | 0.009 |
| DRC | Paid work | ε,EM2 | 0.195 | 0.008 |
| DRC | Paid work | ε,EM1 | 0.212 | 0.008 |
| DRC | Decision ai | vEM3 | 0.244 | 0.038 |
| DRC | Decision ai | βEM,2,1 | 0.397 | 0.041 |
| DRC | Decision ai | λEM,2,1 | -0.007 | 0.041 |
| DRC | Decision ai | λEM,2,1 | -0.01 | 0.039 |

| | | | | |
|---|---|---|---|---|
| DRC | Decision ar | vEM2 | 0.298 | 0.041 |
| DRC | Decision ar | βEM,1,0 | 0.491 | 0.032 |
| DRC | Decision ar | λEM,1,0 | -0.056 | 0.045 |
| DRC | Decision ar | vCT1 | 0.941 | 0.122 |
| DRC | Decision ar | θCT,age | -0.015 | 0.004 |
| DRC | Decision ar | θCT,marria | 0.016 | 0.052 |
| DRC | Decision ar | θCT,highes | 0 | 0.004 |
| DRC | Decision ar | θCT,births | 0.024 | 0.013 |
| DRC | Decision ar | vEM3 | 0.375 | 0.049 |
| DRC | Decision ar | λEM,2,1 | -0.057 | 0.052 |
| DRC | Decision ar | βEM,2,1 | 0.382 | 0.04 |
| DRC | Decision ar | λEM,2,1 | 0.058 | 0.037 |
| DRC | Decision ar | vEM2 | 0.457 | 0.047 |
| DRC | Decision ar | λEM,1,0 | -0.021 | 0.044 |
| DRC | Decision ar | βEM,1,0 | 0.317 | 0.04 |
| DRC | Decision ar | vEM1 | 0.212 | 0.127 |
| DRC | Decision ar | θEM,age | 0.012 | 0.003 |
| DRC | Decision ar | θEM,marria | 0.121 | 0.059 |
| DRC | Decision ar | θEM,highes | 0.005 | 0.002 |
| DRC | Decision ar | θEM,births | -0.03 | 0.01 |
| DRC | Decision ar | cov,CT3,EN | -0.005 | 0.009 |
| DRC | Decision ar | cov,CT2,EN | -0.007 | 0.007 |
| DRC | Decision ar | cov,CT1,EN | -0.008 | 0.01 |
| DRC | Decision ar | ε,EM3 | 0.207 | 0.008 |
| DRC | Decision ar | ε,EM2 | 0.187 | 0.008 |
| DRC | Decision ar | ε,CT1 | 0.24 | 0.006 |
| DRC | Decision ar | ε,EM3 | 0.201 | 0.012 |
| DRC | Decision ar | ε,EM2 | 0.206 | 0.011 |
| DRC | Decision ar | ε,EM1 | 0.225 | 0.008 |
| DRC | Decision ar | vEM3 | 0.256 | 0.053 |
| DRC | Decision ar | βEM,2,1 | 0.403 | 0.041 |
| DRC | Decision ar | λEM,2,1 | -0.052 | 0.06 |
| DRC | Decision ar | λEM,2,1 | 0.022 | 0.048 |
| DRC | Decision ar | vEM2 | 0.347 | 0.06 |
| DRC | Decision ar | βEM,1,0 | 0.496 | 0.032 |
| DRC | Decision ar | λEM,1,0 | -0.107 | 0.066 |
| DRC | Decision ar | vCT1 | 0.936 | 0.122 |
| DRC | Decision ar | θCT,age | -0.014 | 0.004 |
| DRC | Decision ar | θCT,marria | 0.013 | 0.052 |
| DRC | Decision ar | θCT,highes | 0 | 0.004 |
| DRC | Decision ar | θCT,births | 0.024 | 0.013 |
| DRC | Decision ar | vEM3 | 0.562 | 0.055 |
| DRC | Decision ar | λEM,2,1 | -0.02 | 0.05 |
| DRC | Decision ar | βEM,2,1 | 0.319 | 0.071 |

| | | | | |
|---|---|---|---|---|
| DRC | Decision ar | λEM,2,1 | 0.039 | 0.025 |
| DRC | Decision ar | vEM2 | 0.532 | 0.051 |
| DRC | Decision ar | λEM,1,0 | 0.016 | 0.055 |
| DRC | Decision ar | βEM,1,0 | 0.296 | 0.066 |
| DRC | Decision ar | vEM1 | 0.504 | 0.131 |
| DRC | Decision ar | θEM,age | 0.007 | 0.003 |
| DRC | Decision ar | θEM,marria | 0.079 | 0.061 |
| DRC | Decision ar | θEM,highes | 0.001 | 0.002 |
| DRC | Decision ar | θEM,births | 0.002 | 0.008 |
| DRC | Decision ar | cov,CT3,EM | -0.004 | 0.008 |
| DRC | Decision ar | cov,CT2,EM | -0.01 | 0.007 |
| DRC | Decision ar | cov,CT1,EM | 0.003 | 0.011 |
| DRC | Decision ar | ε,EM3 | 0.206 | 0.008 |
| DRC | Decision ar | ε,EM2 | 0.186 | 0.009 |
| DRC | Decision ar | ε,CT1 | 0.241 | 0.006 |
| DRC | Decision ar | ε,EM3 | 0.129 | 0.018 |
| DRC | Decision ar | ε,EM2 | 0.158 | 0.016 |
| DRC | Decision ar | ε,EM1 | 0.149 | 0.019 |
| DRC | Decision ar | vEM3 | 0.219 | 0.023 |
| DRC | Decision ar | βEM,2,1 | 0.43 | 0.03 |
| DRC | Decision ar | λEM,2,1 | -0.019 | 0.041 |
| DRC | Decision ar | λEM,2,1 | 0.023 | 0.043 |
| DRC | Decision ar | vEM2 | 0.23 | 0.027 |
| DRC | Decision ar | βEM,1,0 | 0.533 | 0.027 |
| DRC | Decision ar | λEM,1,0 | 0.006 | 0.038 |
| DRC | Decision ar | vCT1 | 0.368 | 0.074 |
| DRC | Decision ar | θCT,age | -0.001 | 0.003 |
| DRC | Decision ar | θCT,marria | 0.061 | 0.05 |
| DRC | Decision ar | θCT,highes | -0.001 | 0.005 |
| DRC | Decision ar | θCT,births | 0.034 | 0.011 |
| DRC | Decision ar | vEM3 | 0.139 | 0.029 |
| DRC | Decision ar | λEM,2,1 | 0.028 | 0.035 |
| DRC | Decision ar | βEM,2,1 | 0.546 | 0.043 |
| DRC | Decision ar | λEM,2,1 | 0.059 | 0.032 |
| DRC | Decision ar | vEM2 | 0.135 | 0.028 |
| DRC | Decision ar | λEM,1,0 | 0.071 | 0.034 |
| DRC | Decision ar | βEM,1,0 | 0.515 | 0.04 |
| DRC | Decision ar | vEM1 | -0.25 | 0.068 |
| DRC | Decision ar | θEM,age | 0.016 | 0.003 |
| DRC | Decision ar | θEM,marria | 0.159 | 0.044 |
| DRC | Decision ar | θEM,highes | 0.001 | 0.003 |
| DRC | Decision ar | θEM,births | -0.03 | 0.011 |
| DRC | Decision ar | cov,CT3,EM | 0.003 | 0.006 |
| DRC | Decision ar | cov,CT2,EM | 0.003 | 0.007 |

| | | | | |
|---|---|---|---|---|
| DRC | Decision a | cov,CT1,EN | 0.019 | 0.009 |
| DRC | Decision a | ε,EM3 | 0.198 | 0.007 |
| DRC | Decision a | ε,EM2 | 0.179 | 0.007 |
| DRC | Decision a | ε,CT1 | 0.24 | 0.004 |
| DRC | Decision a | ε,EM3 | 0.157 | 0.012 |
| DRC | Decision a | ε,EM2 | 0.153 | 0.012 |
| DRC | Decision a | ε,EM1 | 0.145 | 0.01 |
| DRC | Decision a | vEM3 | 0.219 | 0.044 |
| DRC | Decision a | βEM,2,1 | 0.402 | 0.041 |
| DRC | Decision a | λEM,2,1 | 0.007 | 0.038 |
| DRC | Decision a | λEM,2,1 | 0.019 | 0.05 |
| DRC | Decision a | vEM2 | 0.261 | 0.037 |
| DRC | Decision a | βEM,1,0 | 0.498 | 0.031 |
| DRC | Decision a | λEM,1,0 | 0 | 0.036 |
| DRC | Decision a | vCT1 | 0.949 | 0.119 |
| DRC | Decision a | θCT,age | -0.015 | 0.004 |
| DRC | Decision a | θCT,marria | 0.01 | 0.051 |
| DRC | Decision a | θCT,highes | 0.001 | 0.004 |
| DRC | Decision a | θCT,births | 0.025 | 0.013 |
| DRC | Decision a | vEM3 | 0.508 | 0.057 |
| DRC | Decision a | λEM,2,1 | -0.071 | 0.054 |
| DRC | Decision a | βEM,2,1 | 0.219 | 0.053 |
| DRC | Decision a | λEM,2,1 | 0.014 | 0.042 |
| DRC | Decision a | vEM2 | 0.452 | 0.043 |
| DRC | Decision a | λEM,1,0 | -0.01 | 0.048 |
| DRC | Decision a | βEM,1,0 | 0.281 | 0.05 |
| DRC | Decision a | vEM1 | 0.439 | 0.092 |
| DRC | Decision a | θEM,age | 0.004 | 0.003 |
| DRC | Decision a | θEM,marria | 0.125 | 0.049 |
| DRC | Decision a | θEM,highes | 0.008 | 0.005 |
| DRC | Decision a | θEM,births | -0.024 | 0.012 |
| DRC | Decision a | cov,CT3,EN | -0.016 | 0.009 |
| DRC | Decision a | cov,CT2,EN | -0.011 | 0.007 |
| DRC | Decision a | cov,CT1,EN | -0.003 | 0.011 |
| DRC | Decision a | ε,EM3 | 0.207 | 0.008 |
| DRC | Decision a | ε,EM2 | 0.187 | 0.008 |
| DRC | Decision a | ε,CT1 | 0.24 | 0.006 |
| DRC | Decision a | ε,EM3 | 0.225 | 0.009 |
| DRC | Decision a | ε,EM2 | 0.218 | 0.009 |
| DRC | Decision a | ε,EM1 | 0.235 | 0.006 |
| DRC | Decision a | vEM3 | 0.236 | 0.03 |
| DRC | Decision a | βEM,2,1 | 0.435 | 0.025 |
| DRC | Decision a | λEM,2,1 | -0.03 | 0.023 |
| DRC | Decision a | λEM,2,1 | 0.006 | 0.032 |

| | | | | |
|---|---|---|---|---|
| DRC | Decision ar | vEM2 | 0.254 | 0.031 |
| DRC | Decision ar | βEM,1,0 | 0.515 | 0.024 |
| DRC | Decision ar | λEM,1,0 | -0.018 | 0.03 |
| DRC | Decision ar | vCT1 | 0.442 | 0.066 |
| DRC | Decision ar | θCT,age | -0.004 | 0.003 |
| DRC | Decision ar | θCT,marria | 0.085 | 0.041 |
| DRC | Decision ar | θCT,highes | 0.002 | 0.002 |
| DRC | Decision ar | θCT,births | 0.038 | 0.01 |
| DRC | Decision ar | vEM3 | 0.553 | 0.038 |
| DRC | Decision ar | λEM,2,1 | 0 | 0.032 |
| DRC | Decision ar | βEM,2,1 | 0.321 | 0.046 |
| DRC | Decision ar | λEM,2,1 | -0.025 | 0.03 |
| DRC | Decision ar | vEM2 | 0.5 | 0.044 |
| DRC | Decision ar | λEM,1,0 | -0.04 | 0.028 |
| DRC | Decision ar | βEM,1,0 | 0.25 | 0.046 |
| DRC | Decision ar | vEM1 | 0.805 | 0.054 |
| DRC | Decision ar | θEM,age | -0.002 | 0.002 |
| DRC | Decision ar | θEM,marria | 0.008 | 0.042 |
| DRC | Decision ar | θEM,highes | -0.002 | 0.003 |
| DRC | Decision ar | θEM,births | -0.02 | 0.01 |
| DRC | Decision ar | cov,CT3,EM | 0.003 | 0.006 |
| DRC | Decision ar | cov,CT2,EM | -0.001 | 0.007 |
| DRC | Decision ar | cov,CT1,EM | 0.013 | 0.006 |
| DRC | Decision ar | ε,EM3 | 0.196 | 0.005 |
| DRC | Decision ar | ε,EM2 | 0.184 | 0.006 |
| DRC | Decision ar | ε,CT1 | 0.238 | 0.004 |
| DRC | Decision ar | ε,EM3 | 0.162 | 0.011 |
| DRC | Decision ar | ε,EM2 | 0.212 | 0.011 |
| DRC | Decision ar | ε,EM1 | 0.204 | 0.01 |
| DRC | Decision ar | vEM3 | 0.23 | 0.033 |
| DRC | Decision ar | βEM,2,1 | 0.438 | 0.025 |
| DRC | Decision ar | λEM,2,1 | -0.004 | 0.028 |
| DRC | Decision ar | λEM,2,1 | -0.011 | 0.028 |
| DRC | Decision ar | vEM2 | 0.239 | 0.03 |
| DRC | Decision ar | βEM,1,0 | 0.509 | 0.024 |
| DRC | Decision ar | λEM,1,0 | 0.007 | 0.029 |
| DRC | Decision ar | vCT1 | 0.439 | 0.066 |
| DRC | Decision ar | θCT,age | -0.004 | 0.003 |
| DRC | Decision ar | θCT,marria | 0.075 | 0.04 |
| DRC | Decision ar | θCT,highes | 0.002 | 0.005 |
| DRC | Decision ar | θCT,births | 0.038 | 0.01 |
| DRC | Decision ar | vEM3 | 0.682 | 0.036 |
| DRC | Decision ar | λEM,2,1 | 0.054 | 0.024 |
| DRC | Decision ar | βEM,2,1 | 0.196 | 0.04 |

| | | | | |
|---|---|---|---|---|
| DRC | Decision ai | λEM,2,1 | -0.01 | 0.02 |
| DRC | Decision ai | vEM2 | 0.514 | 0.048 |
| DRC | Decision ai | λEM,1,0 | 0.018 | 0.03 |
| DRC | Decision ai | βEM,1,0 | 0.229 | 0.043 |
| DRC | Decision ai | vEM1 | 0.725 | 0.05 |
| DRC | Decision ai | θEM,age | 0.002 | 0.002 |
| DRC | Decision ai | θEM,marria | 0.023 | 0.043 |
| DRC | Decision ai | θEM,highes | 0.004 | 0.003 |
| DRC | Decision ai | θEM,births | -0.01 | 0.01 |
| DRC | Decision ai | cov,CT3,EM | 0.008 | 0.004 |
| DRC | Decision ai | cov,CT2,EM | 0.009 | 0.007 |
| DRC | Decision ai | cov,CT1,EM | 0.008 | 0.007 |
| DRC | Decision ai | ε,EM3 | 0.197 | 0.005 |
| DRC | Decision ai | ε,EM2 | 0.185 | 0.006 |
| DRC | Decision ai | ε,CT1 | 0.239 | 0.004 |
| DRC | Decision ai | ε,EM3 | 0.126 | 0.012 |
| DRC | Decision ai | ε,EM2 | 0.202 | 0.015 |
| DRC | Decision ai | ε,EM1 | 0.182 | 0.014 |
| Kano | Paid work | vEM3 | 0.141 | 0.032 |
| Kano | Paid work | βEM,2,1 | 0.547 | 0.052 |
| Kano | Paid work | λEM,2,1 | -0.051 | 0.027 |
| Kano | Paid work | λEM,2,1 | 0.021 | 0.032 |
| Kano | Paid work | vEM2 | 0.058 | 0.021 |
| Kano | Paid work | βEM,1,0 | 0.487 | 0.07 |
| Kano | Paid work | λEM,1,0 | 0 | 0.027 |
| Kano | Paid work | vCT1 | -0.156 | 0.071 |
| Kano | Paid work | θCT,age | 0 | 0.002 |
| Kano | Paid work | θCT,marria | 0.127 | 0.036 |
| Kano | Paid work | θCT,highes | 0.056 | 0.021 |
| Kano | Paid work | θCT,births | 0.015 | 0.006 |
| Kano | Paid work | θCT,urban | 0.115 | 0.041 |
| Kano | Paid work | vEM3 | 0.383 | 0.051 |
| Kano | Paid work | λEM,2,1 | 0.105 | 0.046 |
| Kano | Paid work | βEM,2,1 | 0.333 | 0.049 |
| Kano | Paid work | λEM,2,1 | 0.115 | 0.077 |
| Kano | Paid work | vEM2 | 0.43 | 0.038 |
| Kano | Paid work | λEM,1,0 | 0.121 | 0.047 |
| Kano | Paid work | βEM,1,0 | 0.358 | 0.046 |
| Kano | Paid work | vEM1 | 0.058 | 0.113 |
| Kano | Paid work | θEM,age | 0.014 | 0.004 |
| Kano | Paid work | θEM,marria | 0.038 | 0.087 |
| Kano | Paid work | θEM,highes | 0.085 | 0.036 |
| Kano | Paid work | θEM,births | 0.019 | 0.012 |
| Kano | Paid work | θEM,urban | -0.062 | 0.098 |

| | | | | |
|---|---|---|---|---|
| Kano | Paid work | cov,CT3,EM | 0.01 | 0.007 |
| Kano | Paid work | cov,CT2,EM | 0.006 | 0.006 |
| Kano | Paid work | cov,CT1,EM | 0.009 | 0.005 |
| Kano | Paid work | ε,EM3 | 0.118 | 0.016 |
| Kano | Paid work | ε,EM2 | 0.072 | 0.014 |
| Kano | Paid work | ε,CT1 | 0.075 | 0.015 |
| Kano | Paid work | ε,EM3 | 0.205 | 0.012 |
| Kano | Paid work | ε,EM2 | 0.193 | 0.011 |
| Kano | Paid work | ε,EM1 | 0.213 | 0.011 |
| Kano | Decision a | vEM3 | 0.119 | 0.033 |
| Kano | Decision a | βEM,2,1 | 0.492 | 0.054 |
| Kano | Decision a | λEM,2,1 | 0.067 | 0.055 |
| Kano | Decision a | λEM,2,1 | 0.063 | 0.044 |
| Kano | Decision a | vEM2 | 0.059 | 0.023 |
| Kano | Decision a | βEM,1,0 | 0.48 | 0.074 |
| Kano | Decision a | λEM,1,0 | 0.053 | 0.038 |
| Kano | Decision a | vCT1 | 0.571 | 0.428 |
| Kano | Decision a | θCT,age | -0.001 | 0.002 |
| Kano | Decision a | θCT,marria | -0.595 | 0.438 |
| Kano | Decision a | θCT,highes | 0.061 | 0.024 |
| Kano | Decision a | θCT,births | 0.017 | 0.006 |
| Kano | Decision a | θCT,urban | 0.163 | 0.051 |
| Kano | Decision a | vEM3 | 0.193 | 0.061 |
| Kano | Decision a | λEM,2,1 | 0.036 | 0.071 |
| Kano | Decision a | βEM,2,1 | 0.493 | 0.072 |
| Kano | Decision a | λEM,2,1 | 0.179 | 0.052 |
| Kano | Decision a | vEM2 | 0.172 | 0.056 |
| Kano | Decision a | λEM,1,0 | 0.202 | 0.084 |
| Kano | Decision a | βEM,1,0 | 0.371 | 0.068 |
| Kano | Decision a | vEM1 | 0.182 | 0.267 |
| Kano | Decision a | θEM,age | 0 | 0.004 |
| Kano | Decision a | θEM,marria | -0.091 | 0.273 |
| Kano | Decision a | θEM,highes | 0.039 | 0.051 |
| Kano | Decision a | θEM,births | 0.015 | 0.015 |
| Kano | Decision a | θEM,urban | 0.374 | 0.141 |
| Kano | Decision a | cov,CT3,EM | 0.001 | 0.013 |
| Kano | Decision a | cov,CT2,EM | 0.005 | 0.005 |
| Kano | Decision a | cov,CT1,EM | 0 | 0.008 |
| Kano | Decision a | ε,EM3 | 0.14 | 0.018 |
| Kano | Decision a | ε,EM2 | 0.089 | 0.019 |
| Kano | Decision a | ε,CT1 | 0.094 | 0.019 |
| Kano | Decision a | ε,EM3 | 0.171 | 0.027 |
| Kano | Decision a | ε,EM2 | 0.176 | 0.026 |
| Kano | Decision a | ε,EM1 | 0.171 | 0.024 |

| | | | | |
|---|---|---|---|---|
| Kano | Decision ai | vEM3 | 0.119 | 0.038 |
| Kano | Decision ai | βEM,2,1 | 0.493 | 0.058 |
| Kano | Decision ai | λEM,2,1 | 0.096 | 0.069 |
| Kano | Decision ai | λEM,2,1 | -0.013 | 0.058 |
| Kano | Decision ai | vEM2 | 0.06 | 0.024 |
| Kano | Decision ai | βEM,1,0 | 0.478 | 0.077 |
| Kano | Decision ai | λEM,1,0 | 0.044 | 0.038 |
| Kano | Decision ai | vCT1 | 0.577 | 0.427 |
| Kano | Decision ai | θCT,age | -0.001 | 0.002 |
| Kano | Decision ai | θCT,marria | -0.593 | 0.437 |
| Kano | Decision ai | θCT,highes | 0.06 | 0.024 |
| Kano | Decision ai | θCT,births | 0.017 | 0.006 |
| Kano | Decision ai | θCT,urban | 0.168 | 0.049 |
| Kano | Decision ai | vEM3 | 0.219 | 0.066 |
| Kano | Decision ai | λEM,2,1 | 0.054 | 0.073 |
| Kano | Decision ai | βEM,2,1 | 0.567 | 0.086 |
| Kano | Decision ai | λEM,2,1 | 0.065 | 0.067 |
| Kano | Decision ai | vEM2 | 0.27 | 0.081 |
| Kano | Decision ai | λEM,1,0 | 0.148 | 0.089 |
| Kano | Decision ai | βEM,1,0 | 0.433 | 0.09 |
| Kano | Decision ai | vEM1 | 0.098 | 0.283 |
| Kano | Decision ai | θEM,age | -0.002 | 0.004 |
| Kano | Decision ai | θEM,marria | 0.166 | 0.275 |
| Kano | Decision ai | θEM,highes | 0.038 | 0.061 |
| Kano | Decision ai | θEM,births | 0.009 | 0.009 |
| Kano | Decision ai | θEM,urban | 0.531 | 0.114 |
| Kano | Decision ai | cov,CT3,EN | 0.001 | 0.008 |
| Kano | Decision ai | cov,CT2,EN | 0.002 | 0.007 |
| Kano | Decision ai | cov,CT1,EN | -0.006 | 0.007 |
| Kano | Decision ai | ε,EM3 | 0.143 | 0.016 |
| Kano | Decision ai | ε,EM2 | 0.093 | 0.019 |
| Kano | Decision ai | ε,CT1 | 0.096 | 0.019 |
| Kano | Decision ai | ε,EM3 | 0.166 | 0.025 |
| Kano | Decision ai | ε,EM2 | 0.198 | 0.021 |
| Kano | Decision ai | ε,EM1 | 0.179 | 0.021 |
| Kano | Decision ai | vEM3 | 0.139 | 0.032 |
| Kano | Decision ai | βEM,2,1 | 0.509 | 0.054 |
| Kano | Decision ai | λEM,2,1 | -0.046 | 0.028 |
| Kano | Decision ai | λEM,2,1 | 0.07 | 0.034 |
| Kano | Decision ai | vEM2 | 0.06 | 0.023 |
| Kano | Decision ai | βEM,1,0 | 0.489 | 0.075 |
| Kano | Decision ai | λEM,1,0 | 0.021 | 0.03 |
| Kano | Decision ai | vCT1 | -0.203 | 0.094 |
| Kano | Decision ai | θCT,age | 0 | 0.003 |

| | | | | |
|---|---|---|---|---|
| Kano | Decision a | θCT,marria | 0.152 | 0.054 |
| Kano | Decision a | θCT,highes | 0.064 | 0.023 |
| Kano | Decision a | θCT,births | 0.017 | 0.006 |
| Kano | Decision a | θCT,urban | 0.15 | 0.047 |
| Kano | Decision a | vEM3 | 0.289 | 0.059 |
| Kano | Decision a | λEM,2,1 | 0.077 | 0.047 |
| Kano | Decision a | βEM,2,1 | 0.444 | 0.061 |
| Kano | Decision a | λEM,2,1 | 0.125 | 0.087 |
| Kano | Decision a | vEM2 | 0.358 | 0.054 |
| Kano | Decision a | λEM,1,0 | 0.148 | 0.041 |
| Kano | Decision a | βEM,1,0 | 0.381 | 0.075 |
| Kano | Decision a | vEM1 | -0.482 | 0.159 |
| Kano | Decision a | θEM,age | 0.01 | 0.003 |
| Kano | Decision a | θEM,marria | 0.445 | 0.149 |
| Kano | Decision a | θEM,highes | 0.134 | 0.045 |
| Kano | Decision a | θEM,births | 0.03 | 0.007 |
| Kano | Decision a | θEM,urban | 0.104 | 0.12 |
| Kano | Decision a | cov,CT3,EM | 0.006 | 0.008 |
| Kano | Decision a | cov,CT2,EM | 0.007 | 0.007 |
| Kano | Decision a | cov,CT1,EM | 0.014 | 0.006 |
| Kano | Decision a | ε,EM3 | 0.136 | 0.014 |
| Kano | Decision a | ε,EM2 | 0.085 | 0.017 |
| Kano | Decision a | ε,CT1 | 0.09 | 0.017 |
| Kano | Decision a | ε,EM3 | 0.19 | 0.015 |
| Kano | Decision a | ε,EM2 | 0.205 | 0.015 |
| Kano | Decision a | ε,EM1 | 0.196 | 0.011 |
| Kano | Decision a | vEM3 | 0.141 | 0.031 |
| Kano | Decision a | βEM,2,1 | 0.49 | 0.054 |
| Kano | Decision a | λEM,2,1 | 0.039 | 0.055 |
| Kano | Decision a | λEM,2,1 | 0.065 | 0.049 |
| Kano | Decision a | vEM2 | 0.059 | 0.02 |
| Kano | Decision a | βEM,1,0 | 0.473 | 0.074 |
| Kano | Decision a | λEM,1,0 | 0.096 | 0.028 |
| Kano | Decision a | vCT1 | 0.571 | 0.428 |
| Kano | Decision a | θCT,age | -0.001 | 0.002 |
| Kano | Decision a | θCT,marria | -0.594 | 0.437 |
| Kano | Decision a | θCT,highes | 0.061 | 0.024 |
| Kano | Decision a | θCT,births | 0.017 | 0.006 |
| Kano | Decision a | θCT,urban | 0.164 | 0.05 |
| Kano | Decision a | vEM3 | 0.154 | 0.049 |
| Kano | Decision a | λEM,2,1 | 0.039 | 0.071 |
| Kano | Decision a | βEM,2,1 | 0.309 | 0.072 |
| Kano | Decision a | λEM,2,1 | -0.029 | 0.069 |
| Kano | Decision a | vEM2 | 0.124 | 0.042 |

| | | | | |
|---|---|---|---|---|
| Kano | Decision a | λEM,1,0 | 0.07 | 0.046 |
| Kano | Decision a | βEM,1,0 | 0.13 | 0.043 |
| Kano | Decision a | vEM1 | 0.454 | 0.249 |
| Kano | Decision a | θEM,age | -0.001 | 0.003 |
| Kano | Decision a | θEM,marria | -0.357 | 0.276 |
| Kano | Decision a | θEM,highes | 0.131 | 0.044 |
| Kano | Decision a | θEM,births | 0.01 | 0.01 |
| Kano | Decision a | θEM,urban | -0.063 | 0.081 |
| Kano | Decision a | cov,CT3,EN | 0.006 | 0.009 |
| Kano | Decision a | cov,CT2,EN | 0.001 | 0.006 |
| Kano | Decision a | cov,CT1,EN | 0.006 | 0.004 |
| Kano | Decision a | ε,EM3 | 0.144 | 0.017 |
| Kano | Decision a | ε,EM2 | 0.089 | 0.018 |
| Kano | Decision a | ε,CT1 | 0.094 | 0.019 |
| Kano | Decision a | ε,EM3 | 0.15 | 0.029 |
| Kano | Decision a | ε,EM2 | 0.128 | 0.03 |
| Kano | Decision a | ε,EM1 | 0.132 | 0.019 |
| Kano | Decision a | vEM3 | 0.091 | 0.031 |
| Kano | Decision a | βEM,2,1 | 0.501 | 0.055 |
| Kano | Decision a | λEM,2,1 | 0.074 | 0.037 |
| Kano | Decision a | λEM,2,1 | 0.011 | 0.03 |
| Kano | Decision a | vEM2 | 0.041 | 0.024 |
| Kano | Decision a | βEM,1,0 | 0.46 | 0.066 |
| Kano | Decision a | λEM,1,0 | 0.048 | 0.029 |
| Kano | Decision a | vCT1 | -0.159 | 0.074 |
| Kano | Decision a | θCT,age | 0 | 0.002 |
| Kano | Decision a | θCT,marria | 0.128 | 0.036 |
| Kano | Decision a | θCT,highes | 0.059 | 0.022 |
| Kano | Decision a | θCT,births | 0.017 | 0.006 |
| Kano | Decision a | θCT,urban | 0.124 | 0.049 |
| Kano | Decision a | vEM3 | 0.342 | 0.074 |
| Kano | Decision a | λEM,2,1 | 0.031 | 0.042 |
| Kano | Decision a | βEM,2,1 | 0.465 | 0.086 |
| Kano | Decision a | λEM,2,1 | 0.037 | 0.075 |
| Kano | Decision a | vEM2 | 0.36 | 0.075 |
| Kano | Decision a | λEM,1,0 | 0.086 | 0.068 |
| Kano | Decision a | βEM,1,0 | 0.378 | 0.063 |
| Kano | Decision a | vEM1 | 0.528 | 0.151 |
| Kano | Decision a | θEM,age | 0 | 0.004 |
| Kano | Decision a | θEM,marria | -0.153 | 0.06 |
| Kano | Decision a | θEM,highes | 0.104 | 0.043 |
| Kano | Decision a | θEM,births | 0.015 | 0.01 |
| Kano | Decision a | θEM,urban | -0.232 | 0.116 |
| Kano | Decision a | cov,CT3,EN | 0.017 | 0.009 |

| | | | | |
|---|---|---|---|---|
| Kano | Decision ai | cov,CT2,EN | 0.014 | 0.006 |
| Kano | Decision ai | cov,CT1,EN | 0.016 | 0.007 |
| Kano | Decision ai | ε,EM3 | 0.127 | 0.017 |
| Kano | Decision ai | ε,EM2 | 0.077 | 0.015 |
| Kano | Decision ai | ε,CT1 | 0.081 | 0.016 |
| Kano | Decision ai | ε,EM3 | 0.184 | 0.022 |
| Kano | Decision ai | ε,EM2 | 0.211 | 0.012 |
| Kano | Decision ai | ε,EM1 | 0.233 | 0.009 |
| Kano | Decision ai | vEM3 | 0.092 | 0.032 |
| Kano | Decision ai | βEM,2,1 | 0.492 | 0.058 |
| Kano | Decision ai | λEM,2,1 | 0.11 | 0.029 |
| Kano | Decision ai | λEM,2,1 | -0.025 | 0.038 |
| Kano | Decision ai | vEM2 | 0.029 | 0.021 |
| Kano | Decision ai | βEM,1,0 | 0.462 | 0.067 |
| Kano | Decision ai | λEM,1,0 | 0.07 | 0.025 |
| Kano | Decision ai | vCT1 | -0.172 | 0.079 |
| Kano | Decision ai | θCT,age | 0 | 0.002 |
| Kano | Decision ai | θCT,marria | 0.129 | 0.037 |
| Kano | Decision ai | θCT,highes | 0.061 | 0.023 |
| Kano | Decision ai | θCT,births | 0.015 | 0.006 |
| Kano | Decision ai | θCT,urban | 0.133 | 0.049 |
| Kano | Decision ai | vEM3 | 0.356 | 0.062 |
| Kano | Decision ai | λEM,2,1 | 0.099 | 0.046 |
| Kano | Decision ai | βEM,2,1 | 0.353 | 0.065 |
| Kano | Decision ai | λEM,2,1 | 0.107 | 0.074 |
| Kano | Decision ai | vEM2 | 0.367 | 0.055 |
| Kano | Decision ai | λEM,1,0 | 0.138 | 0.068 |
| Kano | Decision ai | βEM,1,0 | 0.311 | 0.065 |
| Kano | Decision ai | vEM1 | 0.578 | 0.139 |
| Kano | Decision ai | θEM,age | -0.003 | 0.004 |
| Kano | Decision ai | θEM,marria | -0.142 | 0.069 |
| Kano | Decision ai | θEM,highes | 0.091 | 0.044 |
| Kano | Decision ai | θEM,births | 0.012 | 0.01 |
| Kano | Decision ai | θEM,urban | 0.025 | 0.129 |
| Kano | Decision ai | cov,CT3,EN | 0.013 | 0.007 |
| Kano | Decision ai | cov,CT2,EN | 0.019 | 0.004 |
| Kano | Decision ai | cov,CT1,EN | 0.017 | 0.009 |
| Kano | Decision ai | ε,EM3 | 0.126 | 0.016 |
| Kano | Decision ai | ε,EM2 | 0.076 | 0.015 |
| Kano | Decision ai | ε,CT1 | 0.081 | 0.017 |
| Kano | Decision ai | ε,EM3 | 0.208 | 0.017 |
| Kano | Decision ai | ε,EM2 | 0.221 | 0.011 |
| Kano | Decision ai | ε,EM1 | 0.236 | 0.008 |
