## Supplementary figures and images for "Untangling Empowerment and Contraceptive Use Longitudinally in Five African Settings"

### appendix figure

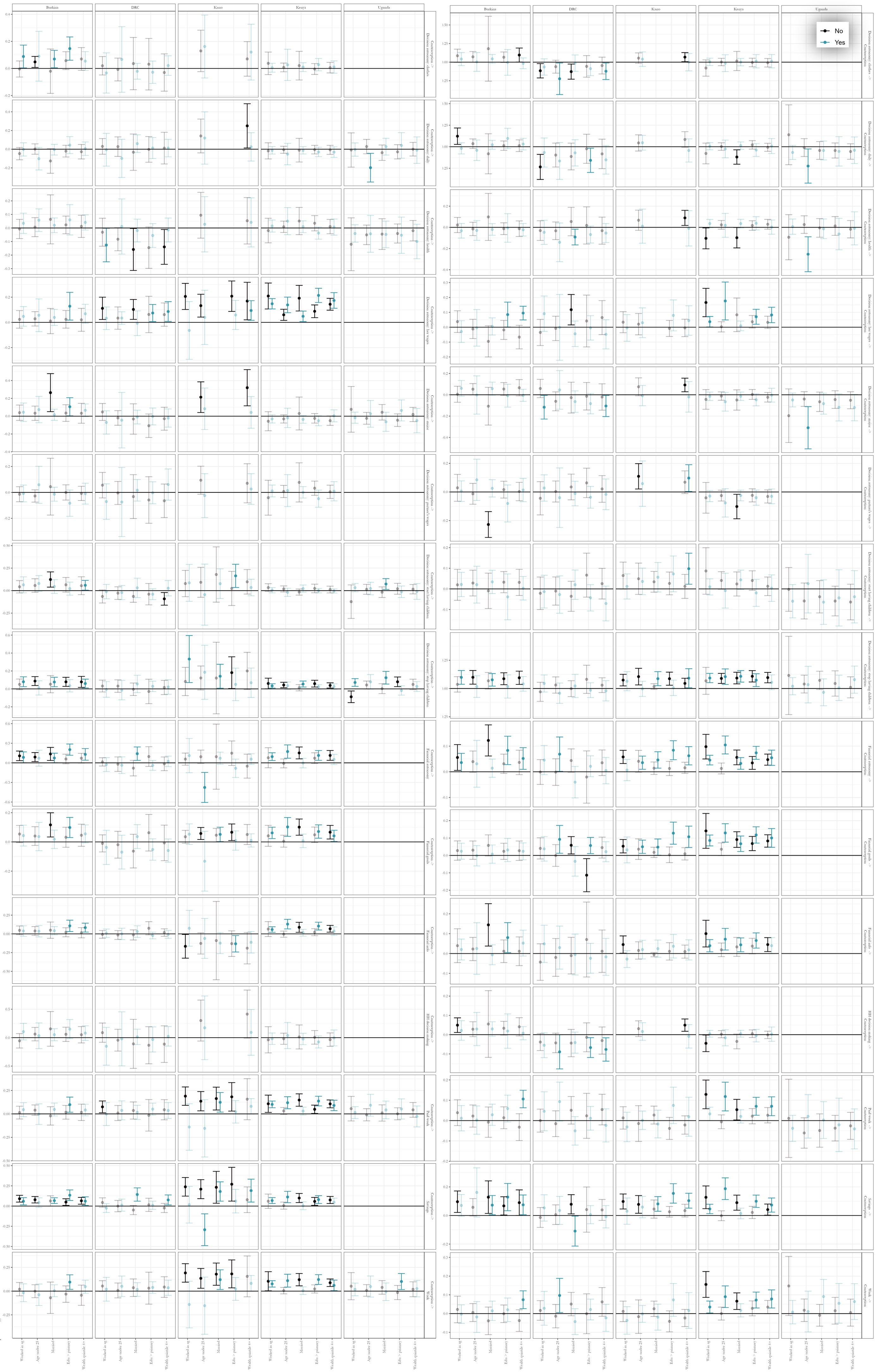
